# Biallelic pathogenic variants in TRMT1 disrupt tRNA modification and induce a syndromic neurodevelopmental disorder

**DOI:** 10.1101/2024.07.18.24310581

**Authors:** Stephanie Efthymiou, Cailyn P Leo, Chenghong Deng, Kejia Zhang, Sheng-Jia Lin, Reza Maroofian, Rauan Kaiyrzhanov, Renee Lin, Irem Karagoz, Annarita Scardamaglia, Daniel Owrang, Valentina Turchetti, Friederike Jahnke, Cassidy Petree, Anna V Derrick, Mark I Rees, Javeria Raza Alvi, Tipu Sultan, Chumei Li, Marie-Line Jacquemont, Frederic Tran-Mau-Them, Maria Valenzuela-Palafoll, Rich Sidlow, Grace Yoon, Michelle Morrow, Alexis Carere, Mary O’Connor, Julie Fleischer, Erica H Gerkes, Chanika Phornphutkul, Bertrand Isidor, Clotilde Rivier-Ringenbach, Christophe Philippe, Semra H Kurul, Didem Soydemir, Bulent Kara, Deniz Sunnetci-Akkoyunlu, Viktoria Bothe, Konrad Platzer, Dagmar Wieczorek, Margarete Koch-Hogrebe, Nils Rahner, Ann-Charlotte Thuresson, Hans Matsson, Carina Frykholm, Sevcan Tuğ Bozdoğan, Atıl Bişgin, Nicolas Chatron, Gaetan Lesca, Sara Cabet, Zeynep Tümer, Tina D Hjortshøj, Gitte Rønde, Thorsten Marquardt, Janine Reunert, Erum Afzal, Mina Zamani, Reza Azizimalamiri, Hamid Galehdari, Pardis Nourbakhshd, Niloofar Chamanrou, Seo-Kyung Chung, Mohnish Suri, Paul J Benke, Maha S Zaki, Joseph G Gleeson, Daniel G Calame, Davut Pehlivan, Halil I Yilmaz, Alper Gezdirici, Aboulfazl Rad, Iman Sabri Abumansour, Gabriela Oprea, Jai Sidpra, Kshitij Mankad, Barbara Vona, Andrew E Fry, Gaurav K Varshney, Henry Houlden, Dragony Fu

## Abstract

The post-transcriptional modification of tRNAs plays a key role in tRNA folding and function to ensure proper levels of protein synthesis during growth and development. Pathogenic variants in tRNA modification enzymes have been implicated in diverse human neurodevelopmental and neurological disorders. However, the molecular basis for many of these disorders remains unknown, thereby limiting our understanding and potential treatment of pathologies linked to tRNA modification. Here, we describe an extensive cohort of 31 individuals from 24 unrelated families with bi-allelic variants in the *tRNA methyltransferase 1* (*TRMT1*) gene who present with a syndromic neurodevelopmental disorder universally characterized by intellectual disability in affected patients. Developmental delay, behavioral abnormalities and facial dysmorphisms represent additional core phenotypes of this syndrome. The variants include novel and ultra-rare *TRMT1* variants that segregate with clinical pathology. We found that a subset of variants causes mis-splicing and loss of TRMT1 protein expression. Notably, patient cells with *TRMT1* variants exhibit a deficiency in tRNA modifications catalyzed by TRMT1. Molecular analysis of *TRMT1* variants reveal distinct regions of the TRMT1 protein required for tRNA modification activity and binding, including a TRMT1 subdomain critical for tRNA interaction. Importantly, depletion of TRMT1 in zebrafish is sufficient to induce developmental and behavioral phenotypes that recapitulate those observed in human patients with pathogenic *TRMT1* variants. Altogether, these findings demonstrate that loss of TRMT1-catalyzed tRNA modifications leads to a syndromic form of intellectual disability and elucidate the molecular underpinnings of tRNA modification deficiency caused by pathogenic TRMT1 variants.

## Introduction

Intellectual disability (ID) is a neurodevelopmental disorder characterized by significant limitations in intellectual ability and adaptive function with a prevalence estimated between 2% and 3% in the general population (1). Genomic sequencing studies have identified an increasing number of causative monogenic variants for ID in genes encoding a diverse group of proteins. Notably, pathogenic variants in genes encoding RNA modification enzymes have been identified as a cause of cognitive disorders in the human population (2–4). These findings highlight the emerging role of tRNA modification in normal neurological development and function.

Human tRNA methyltransferase 1 (TRMT1) is a tRNA modification enzyme that catalyzes the formation of *N*2, *N*2-dimethylguanosine (m2,2G) in cytosolic and mitochondrial tRNAs (5, 6). TRMT1 has been demonstrated to be responsible for nearly all m2,2G modifications in the tRNA of human cells (6, 7). The m2,2G modification has been proposed to play a role in tRNA structure and function (8–10). Human cells deficient in TRMT1 exhibit decreased global protein synthesis and reduced cellular proliferation (6). TRMT1 has also been found to be a cleavage target of the SARS-CoV-2 main protease, suggesting that perturbation of tRNA modification patterns contributes to the cellular pathology of SARS-CoV-2 infection (11, 12). Intriguingly, neuronal activation induces a change in the subcellular distribution of TRMT1, suggesting a role for TRMT1-catalyzed tRNA modification in neuronal transmission and plasticity (7).

Frameshift variants in *TRMT1* have been identified as the cause for certain cases of autosomal-recessive ID through exome sequencing (MRT68, MIM# 618302) (13–16). This was followed by the identification of a single bi-allelic missense variant in *TRMT1* associated with developmental delay, ID, and epilepsy (17). These studies suggest that TRMT1 plays a key role in normal neurodevelopment and cognitive function. However, the impact of *TRMT1* variants on protein expression and function remains unknown for most cases. Moreover, the sparse number of *TRMT1* variants that have been identified and characterized has limited our understanding of cognitive disorders associated with TRMT1 and their physiological consequences.

Here, we describe 31 affected individuals from 24 unrelated families presenting with clinical features of ID in which exome or genome sequencing identified novel and ultra-rare bi-allelic segregating *TRMT1* variants. To functionally characterize the bi-allelic *TRMT1* variants, we explored tRNA modifications in proband-derived fibroblasts or lymphoblasts and quantified m2,2G modifications in cellular tRNAs. Moreover, we investigated the effects of TRMT1-associated ID (TRMT1-ID) variants on reconstitution of activity and interaction between TRMT1 and tRNAs. These studies elucidate the molecular underpinnings of *TRMT1*-derived disorders and significantly expand the spectrum of disease-causing TRMT1 variants.

## Results

### Identification of pathogenic variants in *TRMT1* linked to neurodevelopmental disorders

Using the GeneMatcher platform and data sharing with collaborators, we identified 24 unique families containing 31 individuals affected with neurodevelopmental disorders secondary to bi-allelic variants in *TRMT1* (Figure 1A and 1B, Table 1, Supplementary Table 1). Pedigrees in Figure 1 and Supplementary Table 1 are available upon request from the corresponding authors. We identified 11 missense, 2 nonsense, 13 splice site and 8 frameshift variants. The missense variants were classified as damaging by SIFT, PolyPhen-2, REVEL and Mutation Taster, with a mean CADD score of 27. None of the variants identified were present in the homozygous state in gnomAD v.3.1.2. Nine of the twenty-four identified *TRMT1* variants were absent across multiple genetic databases (∼1 million alleles), whereas the remaining variants appear to be ultra-rare (Supplementary Table 1, available upon request from the corresponding authors).

**Figure 1.**
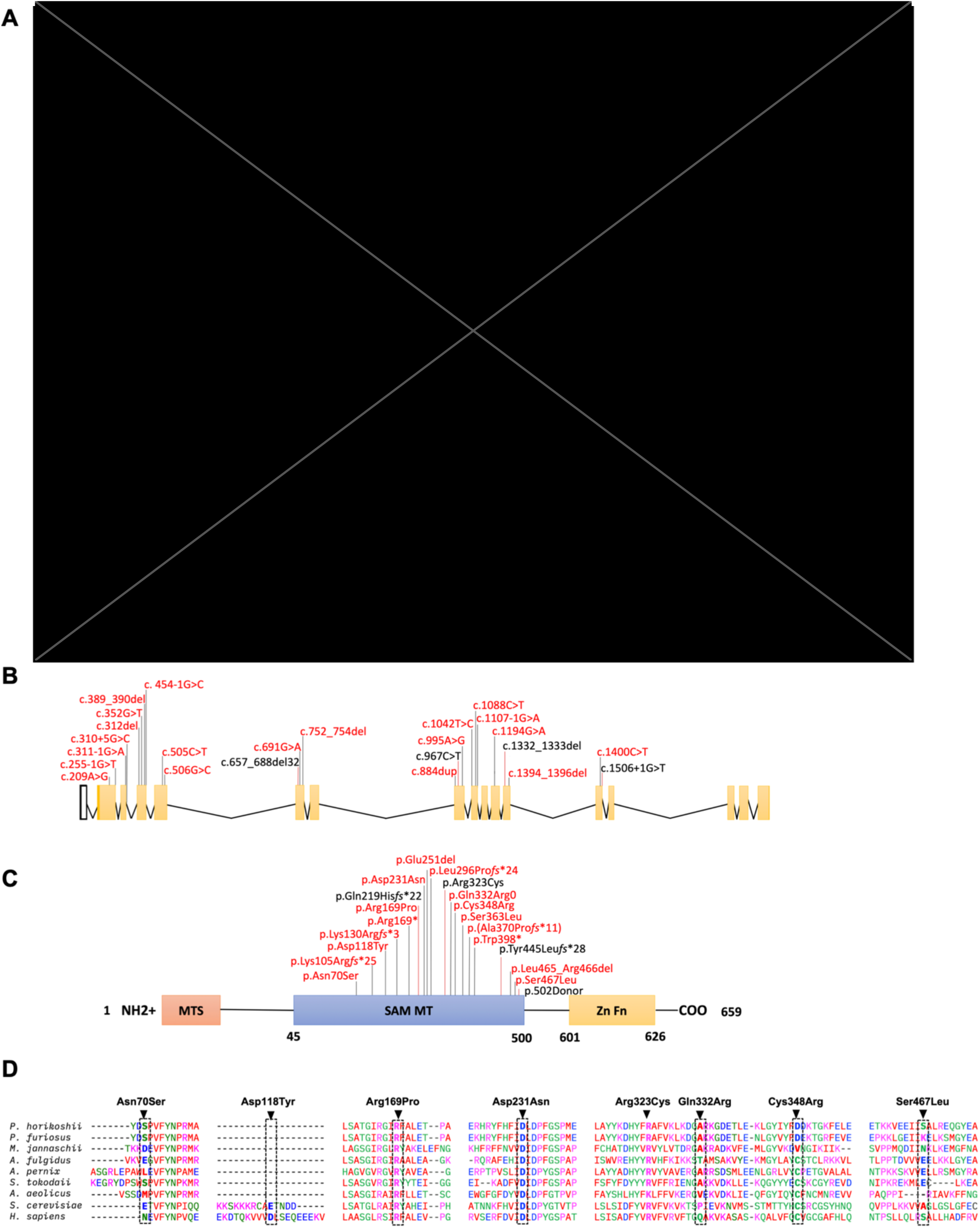
Genetic summary of the reported individuals with homozygous *TRMT1* variants. **(A)** Pedigrees of the 24 families described. Square = male; circle = female; black filled symbol = affected individual; white symbols = unaffected individuals. Double lines indicate consanguinity. **(B)** Coding exons of the *TRMT1* mRNA with variants noted. **(C)** Schematic indicating the domains of the TRMT1 protein. The red box represents the mitochondrial targeting signal while the blue box indicates the class I *S*-adenosyl-methionine (SAM)-dependent methyltransferase domain. The yellow box indicates a C-terminal bipartite nuclear localization signal (NLS) embedded within a C_3_H_1_-type zinc finger motif. Variants reported in this study are represented in red, while previously reported variants in black. **(D)** Protein multiple sequence alignment in *TRMT1* orthologues shows level of conservation of the identified missense residues (indicated in dotted line boxes).

**Table 1.** Clinical Details.

|  | Family 1 |  | Family 2 |  | Family 3 |  |  |
| --- | --- | --- | --- | --- | --- | --- | --- |
| **TRMT1 variant(s)**<br>**(NM_001136035)** | c.454-1G>C p.(Cys86Glnfs*24) |  | c.1194G>A (p.Trp398*) |  | c.1394_1396delTGC (p.Leu465_Arg466delinsTrp) |  |  |
| **Development** |  |  |  |  |  |  |  |
| **Speech/language delay** | Yes | Yes | Yes | Yes | Yes | N/A | N/A |
| **Motor delay** | Yes | Yes | Yes | Yes | Yes | N/A | N/A |
| **ID** | Moderate | Mild | Severe-profound | Mild | Moderate | N/A | N/A |
| **Behaviour** | Yes (poor concentration) | Yes (ADHD) | Yes (Hyperactive and aggressive) | No | No | N/A | N/A |
| **Clinical Features** |  |  |  |  |  |  |  |
| **Microcephaly/Macrocephaly** | No | No | N/A | N/A | No | No | Macrocephaly |
| **Hypertonia or Hypotonia** | - | - | - | - | - | N/A | N/A |
| **Seizures** | - | - | Yes (febrile) | Yes (generalized tonic-clonic) | - | Yes | - |
| **Dysmorphic features** | + | + | + | + | - | N/A | + |
| **Investigations** |  |  |  |  |  |  |  |
| **Abnormal MRI brain** | - | - | + | + | - | N/A | N/A |
| **Abnormal EEG** | + | + | - | - | N/A | N/A | N/A |

Table 1. (Continued)
|  | Family 4 | Family 5 | Family 6 | Family 7 | Family 8 |
| --- | --- | --- | --- | --- | --- |
| **TRMT1 variant(s)**<br>**(NM_001136035)** | c.995A>G<br>(p.Gln332Arg) and<br>c.255-1G>T<br>p.(Cys86Glnfs*24) | c.1107-1G>A<br>p.(Ala370Profs*11) and<br>c.310+5G>C<br>p.(Cys86Glnfs*24) | c.884dup<br>(p.Leu296Profs*24) and<br>c.1042T>C<br>(p.Cys348Arg) | c.209 A>G (p.Asn70Ser)<br>and c.352G>T<br>(p.Asp118Tyr) | c.1107-1G>A<br>p.(Ala370Profs*11) and<br>c.389_390delAA<br>(p.Lys130Argfs*3) |
| **Development** |  |  |  |  |  |
| **Speech/language delay** | Yes | Yes | Yes | Yes | Yes |
| **Motor delay** | Yes | Yes | Yes | No | Yes |
| **ID** | Moderate | Yes | Moderate | Yes | N/A |
| **Behaviour** | Yes (ASD) | No | Yes (ADHD) | Yes (aggressive and non-social) | Yes (Hyperactive) |
| **Clinical Features** |  |  |  |  |  |
| **Microcephaly /Macrocephaly** | Microcephaly | No | No | Macrocephaly | No |
| **Hypertonia or Hypotonia** | - | Hypotonia | - | - | Hypertonia |
| **Seizures** | - | Yes (febrile) | Yes (focal) | - | Yes (generalised) |
| **Dysmorphic features** | + | + | + | + | + |
| **Investigations** |  |  |  |  |  |
| **Abnormal MRI brain** | - | + | + | N/A | + |
| **Abnormal EEG** | - | + | + | N/A | + |

Table 1. (Continued)
|  | Family 9 | Family 10 | Family 11 | Family 12 |
| --- | --- | --- | --- | --- |
| **TRMT1 variant(s)**<br>**(NM_001136035)** | c.506G>C (p.Arg169Pro) | c.310+5G>C p.(Cys86Glnfs*24)<br>and c.967C>T (p.Arg323Cys) | c.505C>T (p.Arg169*) | c.1332_1333del<br>(p.Tyr445Leufs*28) |
| **Development** |  |  |  |  |
| **Speech/language delay** | Yes | Yes | Yes | Yes |
| **Motor delay** | Yes | N/A | N/A | Yes |
| **ID** | Yes | N/A | Yes | Moderate-severe |
| **Behaviour** | N/A | Yes (Hyperactive) | Yes | Yes (Poor concentration) |
| **Clinical Features** |  |  |  |  |
| **Microcephaly /Macrocephaly** | No | No | Microcephaly | Microcephaly |
| **Hypertonia or Hypotonia** | - | N/A | - | Hypotonia |
| **Seizures** | Yes | Yes (generalised febrile) | Yes (febrile) | - |
| **Dysmorphic features** | + | + | + | + |
| **Investigations** |  |  |  |  |
| **Abnormal MRI brain** | - | - | + | + |
| **Abnormal EEG** | N/A | N/A | - | - |

Table 1. (Continued)
|  | Family 13 | Family 14 | Family 15 | Family 16 |
| --- | --- | --- | --- | --- |
| TRMT1 variant(s)<br>(NM_001136035) | c.312del (p.Lys105Argfs*25) | c.691G>A (p.Asp231Asn) | c.1332_1333del<br>(p.Tyr445Leufs*28) | c.311-1G>A p.(Cys86Glnfs*24) |
| Development |  |  |  |  |
| Speech/language delay | Yes | Yes | Yes | Yes |
| Motor delay | Yes | Yes | Yes | Yes |
| ID | Moderate | Moderate | Moderate | Moderate-Severe |
| Behaviour | Yes (Anxiety) | Yes (Hyperactive and self<br>mutilation under-stress) | Yes (Restless, poor autonomy<br>in activities/games) | Yes (ADHD) |
| Clinical Features |  |  |  |  |
| Microcephaly /Macrocephaly | No | No | Microcephaly | Microcephaly |
| Hypertonia or Hypotonia | - | - | Hypotonia | Hypertonia |
| Seizures | Yes (febrile) | Yes (febrile & tonic-clonic) | - | Yes (focal & febrile) |
| Dysmorphic features | + | + | + | + |
| Investigations |  |  |  |  |
| Abnormal MRI brain | - | - | + | + |
| Abnormal EEG | - | + | + | + |

Table 1. (Continued)
|  | Family 17 | Family 18 | Family 19 | Family 20 |  |
| --- | --- | --- | --- | --- | --- |
| TRMT1 variant(s)<br>(NM_001136035) | c.752_754del<br>(p.Glu251del) | c.310+5G>C<br>p.(Cys86Glnfs*24) | c.1400C>T (p.Ser467Leu) and<br>c.967C>T (p.Arg323Cys) | c.657_688del (p.Gln219Hisfs*22) |  |
| Development |  |  |  |  |  |
| Speech/language delay | Yes | Yes | Yes | Yes | Yes |
| Motor delay | Yes | Yes | Yes | Yes | Yes |
| ID | Mild-moderate | Moderate | Mild | Moderate | Moderate |
| Behaviour | No | No | No | No | No |
| Clinical Features |  |  |  |  |  |
| Microcephaly /Macrocephaly | No | No | Microcephaly | Microcephaly | Microcephaly |
| Hypertonia or Hypotonia | N/A | - | Hypotonia | - | - |
| Seizures | - | - | - | Yes (GTC) | Yes (focal &<br>generalised) |
| Dysmorphic features | + | + | + | + | + |
| Investigations |  |  |  |  |  |
| Abnormal MRI brain | + | - | - | + | N/A |
| Abnormal EEG | - | N/A | - | + | - |

Table 1. (Continued)
|  | Family 21 |  | Family 22 | Family 23 |  | Family 24 |
| --- | --- | --- | --- | --- | --- | --- |
| TRMT1 variant(s)<br>(NM_001136035) | c.657_688del (p.Gln219Hisfs*22) |  | c.657_688del<br>p.Gln219Hisfs*22 | c.1332_1333del (p.Tyr445Leufs*28) |  | c.1332_1333del<br>(p.Tyr445Leufs*28) |
| Development |  |  |  |  |  |  |
| Speech/language delay | Yes | Yes | Yes | Yes | Yes | Yes |
| Motor delay | Yes | Yes | Yes | No | No | Yes |
| ID | Moderate | Mild-moderate | Moderate | Moderate-severe | Severe | Yes |
| Behaviour | Yes (Bad temper) | Yes (Bad temper) | Yes (Poor concentration,<br>hyperactivity) | Yes (Bad temper,<br>self-mutilation) | Yes (Anxiety) | Yes (Moderate attention<br>deficit, anxiety) |
| Clinical Features |  |  |  |  |  |  |
| Microcephaly /Macrocephaly | No | No | Microcephaly | No | No | N/A |
| Hypertonia or Hypotonia | - | - | Hypotonia | - | - | - |
| Seizures | Yes | Yes | - | Yes<br>(febrile &<br>generalised) | Yes<br>(generalised) | - |
| Dysmorphic features | + | + | + | + | + | + |
| Investigations |  |  |  |  |  |  |
| Abnormal MRI brain | + | N/A | + | - | - | N/A |
| Abnormal EEG | - | - | - | - | - | - |

Table 1. (Continued)
|  | Family M300<br>(Najmabadi *et al.*, 2011) |  | Family 9000114<br>(Davarniya *et al.*, 2014) |  |  | Family 1<br>(Blaesius *et al.*, 2018) | Family 2<br>(Blaesius *et al.*, 2018) |  |  | Zhang *et al.*, 2020 |
| --- | --- | --- | --- | --- | --- | --- | --- | --- | --- | --- |
| **TRMT1 variant(s)**<br>**(NM_001136035)** | c.657_688 del32<br>p.Gln219Hisfs*22 |  | c.1332_1333 delGT<br>p.Tr445Leufs*28 |  |  | c.657_688del32<br>p.Q219Hfs*22 | c.1506+1G>T<br>p.502 Donor |  |  | c.967C>T<br>p.Arg323Cys |
| **Development** |  |  |  |  |  |  |  |  |  |  |
| **Speech/language delay** | No | No | Yes | Yes | Yes | Yes | No | No | No | N/A |
| **Motor delay** | Yes | Yes | Yes | Yes | Yes | Yes | No | No | No | No |
| **ID** | Moderate | Moderate | Moderate | Mild | Moderate | Moderate | Moderate | Moderate | Moderate | Yes |
| **Behaviour** | N/A | N/A | N/A | N/A | N/A | N/A | N/A | N/A | N/A | N/A |
| **Clinical Features** |  |  |  |  |  |  |  |  |  |  |
| **Microcephaly**<br>**/Macrocephaly** | No | No | No | No | No | Microcephaly | No | Microcephaly | Microcephaly | No |
| **Hypertonia or Hypotonia** | N/A | N/A | N/A | N/A | N/A | N/A | N/A | N/A | N/A | N/A |
| **Seizures** | No | No | N/A | N/A | N/A | Yes (generalised) | Yes | Yes | Yes | Yes(generalised) |
| **Dysmorphic features** | + | + | + | + | + | - | - | - | - | + |
| **Investigations** |  |  |  |  |  |  |  |  |  |  |
| **Abnormal MRI brain** | + | + | - | - | - | + | N/A | N/A | N/A | No |
| **Abnormal EEG** | N/A | N/A | N/A | N/A | N/A | N/A | N/A | N/A | N/A | N/A |

| Abbreviations |  |
| --- | --- |
| **ADHD** | Attention deficit hyperactivity disorder |
| **ASD** | Autism spectrum disorder |
| **F** | Female |
| **GTC** | Generalised tonic-clonic |
| **M** | Male |
| **m** | Months |
| **N/A** | Not available |
| **y** | Years |

All detected variants were located within the conserved *S*-adenosylmethionine-dependent methyltransferase domain (Figure 1C). Of the missense variants, p.Arg169Pro, p.Asp231Asn and p.Arg323Cys are conserved from yeast to humans, while p.Gln332Arg, p.Cys348Arg and p.Ser467Leu are semi-conserved (Figure 1D). p.Tyr445Leu*fs*\*28 was found in two independent individuals (F12:S1 and F15:S1). Similarly, p.Gln219His*fs\**22 was found in two independent families. Overall, these recurring variants suggest a possible founder effect.

Among the 31 affected individuals, 14 patients were identified with homozygous variants in TRMT1 and 14 patients contained compound heterozygous variants in *TRMT1*. Nineteen of the individuals were male (61%) and 12 were female (39%). Consanguinity was reported in 13 families (54%). Additionally, 11 of the families had a positive family history for neurological diseases (48%). The median age at last follow-up was 11 years. The ethnic composition of the cohort is diverse, including families of Pakistani, Caucasian, Middle Eastern, European, and Hispanic origin (Table 1).

### Patients with bi-allelic TRMT1 variants present with a core set of phenotypic features

The 31 affected individuals with biallelic TRMT1 variants exhibited a core set of phenotypic features encompassing intellectual disability, global developmental delay, and facial dysmorphism (Figure 2A, summarized in Table 1). Video recordings are available for affected individuals from Family 1 (Supplementary Video 1 and 2). Detailed clinical history, case reports, and videos are available upon request from the corresponding authors.

**Figure 2.**
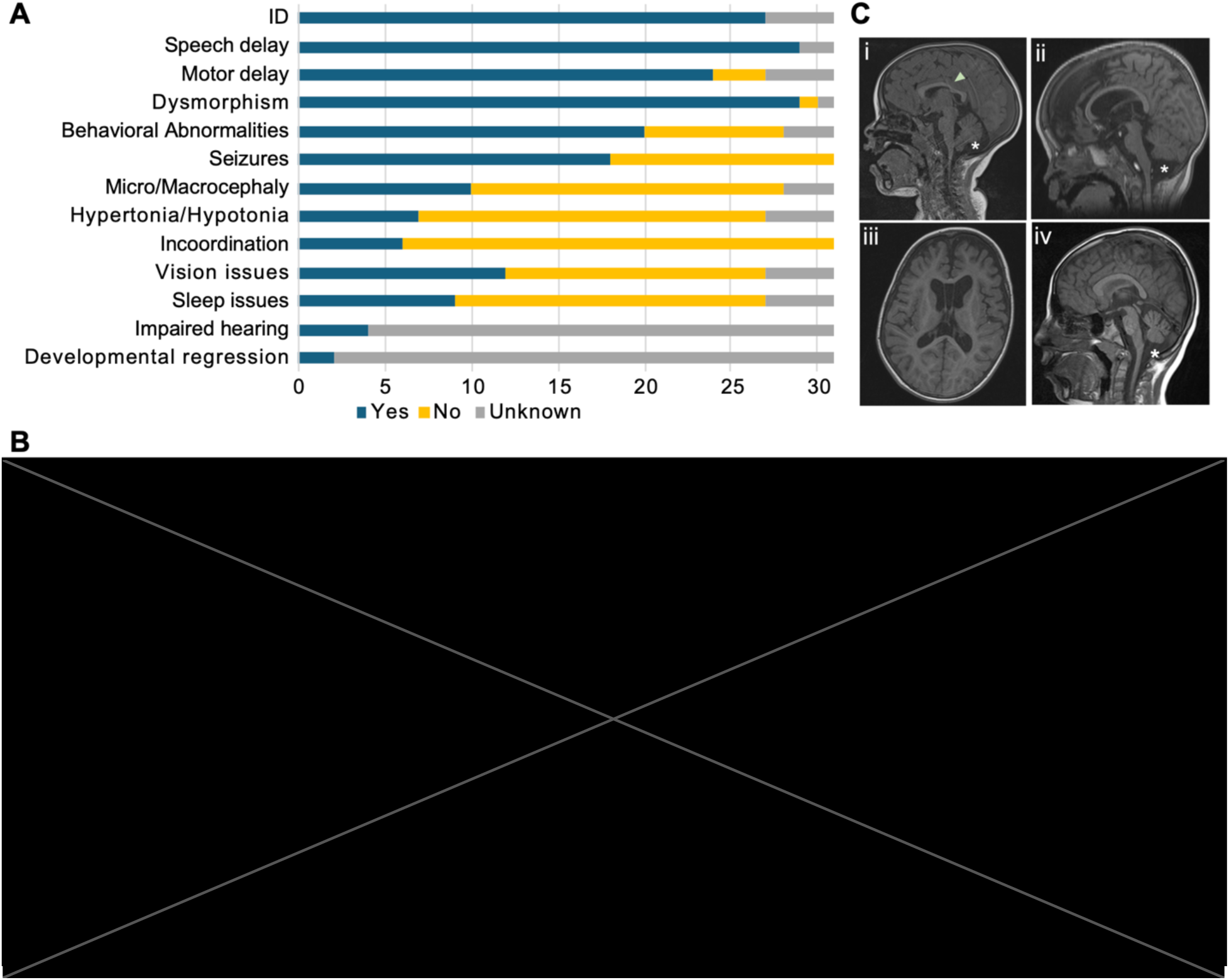
Phenotypic summary of the reported individuals with homozygous *TRMT1* variants. **(A)** Clinical features of the affected individuals with bi-allelic *TRMT1* variants (ID: intellectual disability). **(B)** Frontal facial photographs of *TRMT1* probands showing the most prominent and frequent dysmorphic features of *TRMT1*-related neurodevelopmental delay. **(C)** Representative neuroimaging features identified in individuals with *TRMT1*-ID. (i) Midsagittal T1-weighted MRI of the brain in a childhood boy (F-5) exhibits global (cerebral and cerebellar) atrophy, posterior thinning of the corpus callosum (arrow), and a mega cisterna magna (asterisk). (ii-iii) Midsagittal (ii) and axial (iii) T1-weighted MRI of the brain in a childhood boy (F-8) shows further, characteristic features of TRMT1-ID, namely: frontotemporal-predominant cerebral and midbrain atrophy with corresponding ventriculomegaly and uniform thinning of the corpus callosum (not all shown). Note is also made of the right posterior positional plagiocephaly, (iv) Midsagittal T1-weighted brain MRI of childhood-old boy (F-23), exhibits cerebellar atrophy, a mega cisterna magna (asterisk), and downsloping of the corpus callosum.

Intellectual disability (ID) or developmental delay (DD) for individuals of less than 5 years of age was reported in all patients who were tested (27/27). DD/ID varied in severity among individuals and assessed as mild to moderate DD/ID in 17 out of 22 individuals, moderate to severe DD/ID in 4 out of 22, and severe to profound DD/ID in 1 out of 22. Speech and language development was delayed among all participants tested (29/29), with the median age for first words spoken recorded at 40.5 months (range: 17 months to 8 years).

Motor milestones were delayed in the majority of tested cases (24/27), with median ages of 9 months for unsupported sitting and 23 months for independent walking. Other gross motor manifestations included an unsteady/broad-based gait (n=2), clumsiness (n=7), poor coordination, and ataxia (n=6). Seizures were found in 58% of the patients (18/31), with a median onset age of 16.5 months (range: 1 month to 27 months). Seizure semiology varied and included febrile seizures (n=8), focal fits (n=3), and generalized tonic-clonic seizures (n=7). 35% (n=9/24) of the conducted EEGs were reported as abnormal for these patients. Nine of 28 study subjects had microcephaly with one person having developed secondary microcephaly with time (F19). Additionally, two individuals showed macrocephaly.

Facial photographs and/or videos were reviewed for 13 patients from 10 families (Figure 2B, Supplementary Table 2, feature frequencies tabulated in Supplementary Table 3, data available upon request from the corresponding authors). Based on this assessment, the most frequently seen facial dysmorphic features of *TRMT1*-related neurodevelopmental delay include high anterior hairline (46%), narrow forehead/bifrontal/bitemporal narrowing (54%), sparse eyebrows or laterally sparse eyebrows (62%), up-slanting palpebral fissures (46%), wide/broad nasal bridge (54%) and full or broad nasal tip (85%). The facial features found are relatively non-specific and recognizable facial gestalt for this disorder was not appreciated.

Neurological assessments revealed hypotonia in 5 out of 27 individuals (19%), with 2 individuals presenting it as the primary complaint, while hypertonia was observed in 2 individuals. Other neurological features observed in the cohort included poor coordination/ ataxia (6/28) and intention tremor (3/28). Abnormal movements were reported in 9 out of 30 patients, with motor or verbal tics being the most common manifestation (6/9). A diverse range of behavioral issues were reported in 20 out of 28 individuals, ranging from diagnosed ASD and ADHD to parent-reported concerns such as hyperactivity, aggression, anxious behavior, restlessness, poor autonomy, and irritability.

Feeding difficulties affected 41% of the patients (14 out of 29). Sleep apnea was present in 2 patients, with one utilizing CPAP. Additionally, four individuals had hearing impairment.

Brain MRI was available for 12 individuals (Figure 2C, summarized in Supplementary Figure 1). The most prevalent neuroimaging findings in our cohort were cerebral atrophy (7/12; 58%); cerebellar atrophy (6/12; 50%), which was either global (n=2), limited to the vermis (n=2), or limited to the cerebellar hemispheres (n=2); and posterior thinning of the corpus callosum (5/12; 42%). Two individuals (Family 5 and Family 20 Proband 1) exhibited global brain atrophy. Altogether, these findings identify a core syndromic pattern associated with biallelic TRMT1 variants that can co-occur with a diversity of dysmorphic, neurological, and behavioural phenotypes.

### TRMT1 splice site variants alter splicing patterns

A subset of *TRMT1* variants are predicted to alter mRNA splicing patterns based upon *in silico* splice site prediction algorithms (Supplementary Table 4). To test the effects of the TRMT1 variants on splicing, we generated mini-gene splicing reporter plasmids cloned from the genomic DNA of a healthy wildtype (WT) donor or affected patients. The splicing reporters were transfected into 293T human embryonic cells and splicing analyzed by RT-PCR, sequencing, and fragment analysis (Supplementary Figure 2, Supplementary Data 1, Supplementary Table 5 and Supplementary Table 6).

The c.255-1G>T and c.310+5G>C variants are predicted to abolish the splice acceptor and donor sites of exon 3, respectively, while the c.311-1G>A variant is predicted to abolish the splice acceptor site of exon 4. The c.454-1G>C variant is predicted to eliminate the splice acceptor site of exon 5. The c.255-1G>T, c.310+5G>C, c.311-1G>A, and c.454-1G>C variants were tested using a construct containing introns 2 to 5. RT-PCR spanning exons 3 through 5 from cells transfected with the WT construct showed a complex splicing pattern due to alternative splicing that was analyzed through Sanger sequencing and fragment analysis (Figure 3A, Supplementary Data 1). From fragment analysis, a total of 53% of protein coding transcripts in WT include exons 3 to 5 that were completely eliminated in assays for the c.255-1G>T, c.310+5G>C, and c.454-1G>C variants with an abundance of only 10.6% in sample c.311-1G>A (r.255_641del, p.Cys86Gln*fs\**24) (Supplementary Table 5 and Supplementary Table 6).

**Figure 3.**
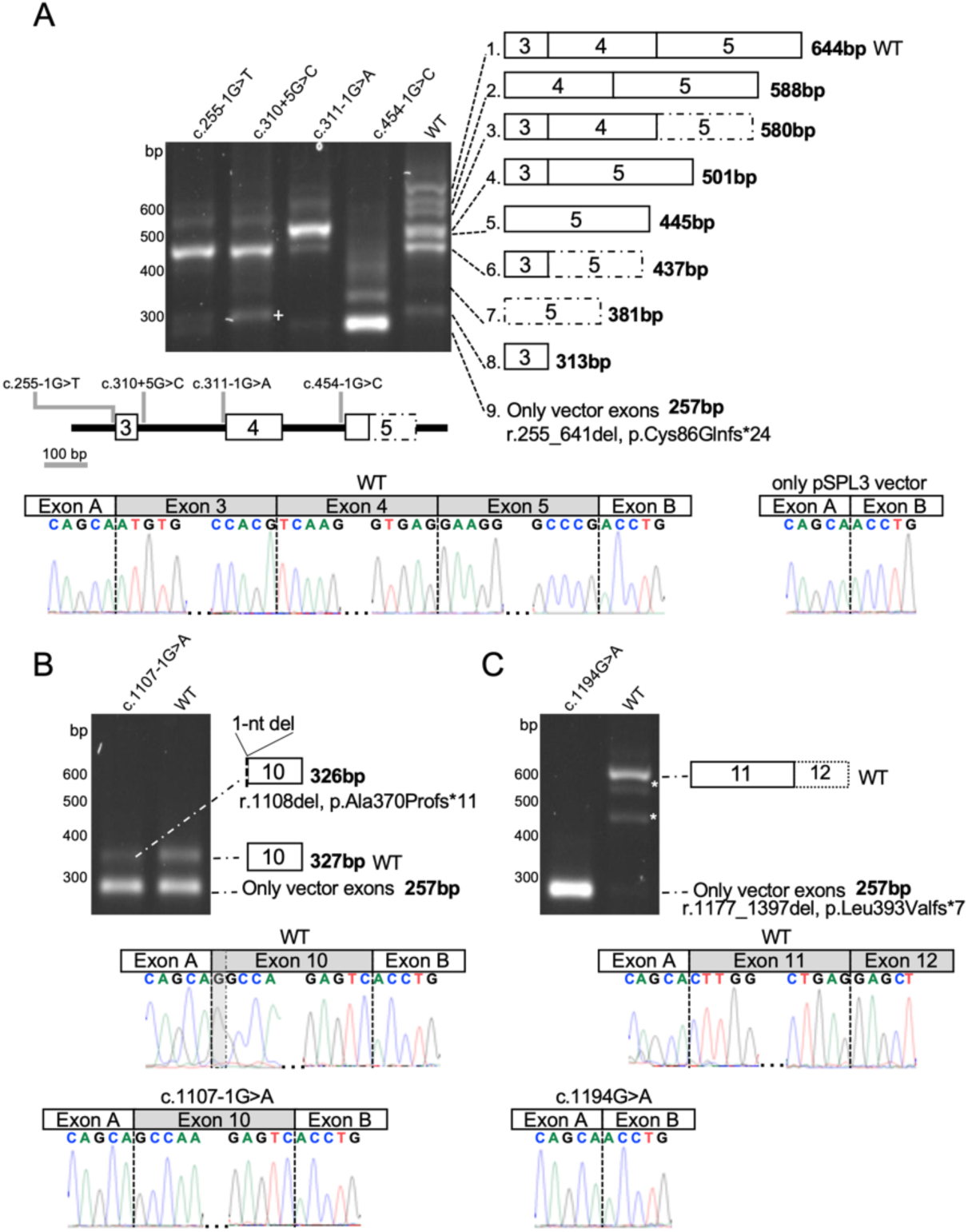
*TRMT1* variants induce splicing defects. Wild-type (WT) and mutant minigenes were transfected in HEK293T cells and their RNA subjected to RT-PCR. **(A)** RT-PCR for WT and mutant minigenes showed complex splicing patterns. We obtained sequencing validation for all except amplicon 2. The presence of additional bands in the WT is attributed to alternative splicing and was quantified in Supplementary Table 5. The only protein coding transcript expressed in the brain includes exons 3-5 that has been eliminated or reduced due to the four variants. The upper left panel shows the RT-PCR agarose gel with the splicing schematic shown for each band to the right. The variant schematic is shown below. Sanger sequencing results of the RT-PCR showing the correctly spliced WT with the deleterious variant effect, exon skipping, as shown by the presence of only pSPL3 vector. The dotted box represents the short version of exon 5. **(B)** RT-PCR of the c.1107-1G>A results in loss of the native splice acceptor site and cryptic splice activation leading to loss of a G and a frameshift. The left panel shows the RT-PCR agarose gel with the splicing schematic shown for each band to the right. Exon 10 with 1 bp deletion is represented with a dotted line corresponding to the position of the single bp deletion. **(C)** RT-PCR of the c.1194G>A variant leads to a frameshift due to exon skipping of exons 11 and 12 leading to a frameshift. Asterisks represent assay artifacts. The left panel shows the RT-PCR gel with the splicing schematic shown to the right. Assay design captured part of exon 12 that was correctly spliced in the WT control.

The c.1107-1G>A variant was predicted to abolish the splice acceptor site of exon 10 and create a cryptic splice site 1 nucleotide downstream from the native canonical splice site that likely causes a deletion of 1 bp and a frameshift r.1108del p.(Ala370Profs*11). The c.1107-1G>A variant was evaluated using a construct spanning introns 8 to 10. RT-PCR of both WT and the c.1107-1G>A variant showed two bands which we attribute to the pSPL3 vector (Figure 3B, gel). The frameshift was validated by Sanger sequencing (Figure 3B, chromatograms).

The splice prediction scores for the c.1194G>A variant suggested either a cryptic donor gain 14 bp from the native splice acceptor site or no splice effect. The c.1194G>A variant was assayed using a construct spanning intron 10 to exon 12. The RT-PCR for the c.1194G>A variant showed skipping of exon 11 and 12 leading to a frameshift (r.1177_1397del, p.Leu393Valfs*7) that was validated with Sanger sequencing (Figure 3C). Altogether, our splicing analyses reveal that a subset of *TRMT1* variants can induce aberrant splicing that are expected to reduce mRNA abundance and/or produce altered protein products.

### *TRMT1* variants differentially impact TRMT1 protein levels

To examine the impact of ID-associated *TRMT1* variants, we next investigated TRMT1 protein expression in available patient cell lines using immunoblotting. For the Asp231Asn (D231N) missense variant in Family 14, we obtained fibroblast cells from the heterozygous father (Patient 14f) and the homozygous offspring that were compared to a control fibroblast cell line (control 1, WT fibroblast). The heterozygous D231N fibroblasts exhibited a ∼2-fold increase in TRMT1 protein compared to WT fibroblast cells (Figure 4A, compare lanes 1 and 2; quantified in Figure 4B). The homozygous D231N fibroblast cell line exhibited an even greater ∼5-fold increase in TRMT1 protein levels compared to WT fibroblast cells (Figure 4A, compare lanes 1 and 3; quantified in Figure 4B). These results suggest that the D231N variant affects the folding of TRMT1 leading to increased stability of TRMT1 against degradation and/or turnover.

**Figure 4.**
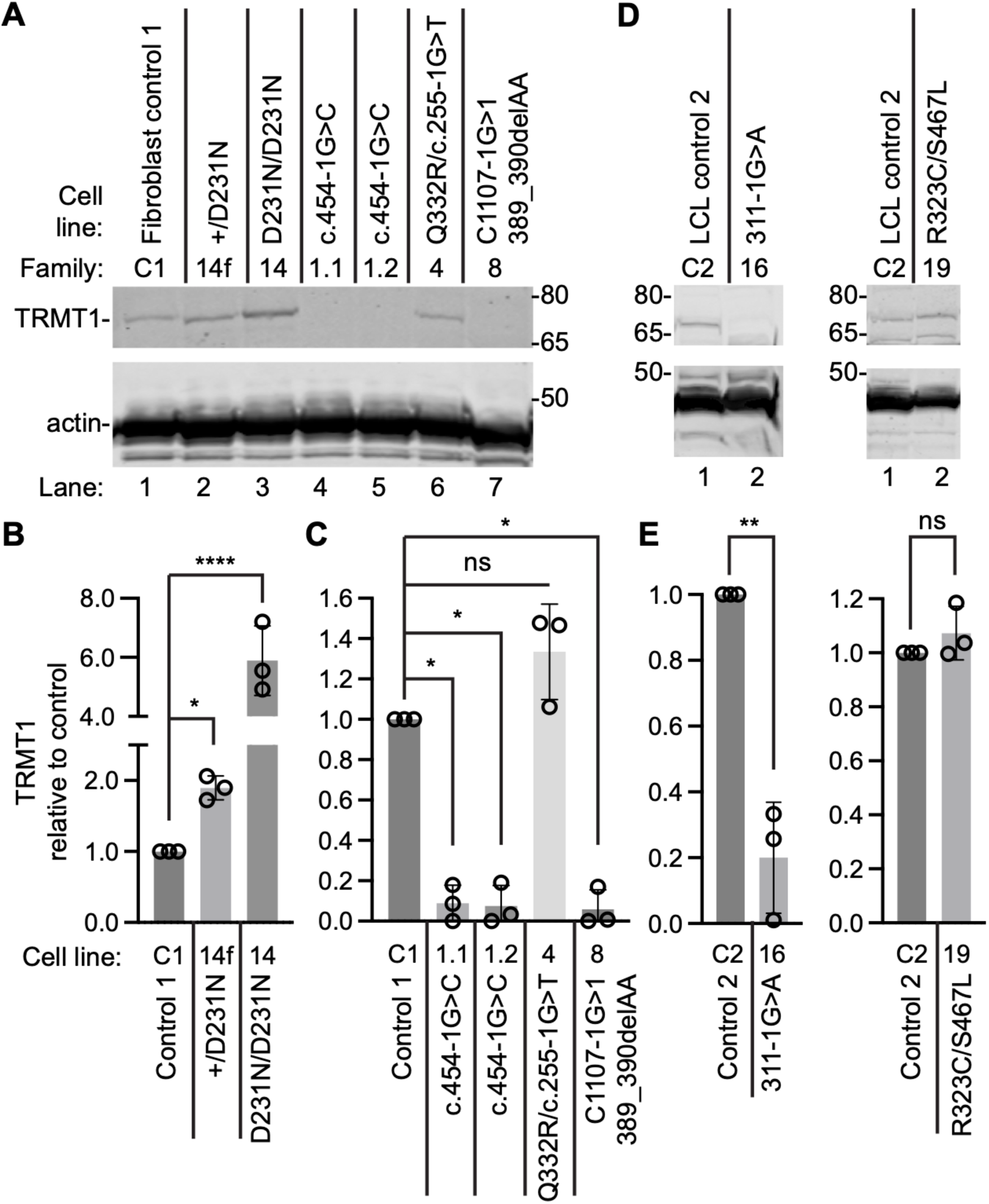
*TRMT1* gene variants cause changes in TRMT1 protein levels. (**A**) Representative immunoblot of the indicated proteins from fibroblast cell lines derived from control (C1) or affected patients. (**B, C**) Quantification of TRMT1 levels relative to the control fibroblast cell line after normalization to actin. N=3. (**D**) Representative immunoblot of TRMT1 and actin from lymphoblast cell lines (LCLs) derived from control wildtype (C2) or affected patients. (**E**) Quantification of TRMT1 levels in LCLs after normalization to actin. N=3. Error bars represent standard deviation from the mean. Statistical analysis was performed using one-way ANOVA. *P ≤ 0.05; **P ≤ 0.01; ***P ≤ 0.001; ****P < 0.0001; ns, non-significant, P > 0.05.

We also generated fibroblast cell lines from patients in families 1, 4, and 8 that harbor *TRMT1* splicing variants (Table 2). We detected nearly complete loss of TRMT1 protein expression in cell lines derived from two different members of Family 1 that have a homozygous splicing variant that eliminates the splice acceptor site of exon 5 (Figure 4A, patients 1.1 and 1.2, lanes 4 and 5; quantified in Figure 4C). Family 8 is compound heterozygous for a splice site and a frameshift variant *in trans*. TRMT1 levels are also reduced to nearly undetectable levels in fibroblasts from Family 8 (Figure 4A, Patient 8, lane 7; quantified in Figure 4C). The fibroblasts from Patient 4 are compound heterozygous for the Gln332Arg (Q332R) missense variant and a splice variant that is predicted to abolish the splice acceptor of exon 3 (c255-1G>T). We did not detect a significant change in TRMT1 protein levels in the fibroblasts from Patient 4 compared to conrols (Figure 4C patient 4, lane 6, quantified in 4C).

For individuals from families 16 and 19, we derived lymphoblastoid cell lines (LCLs) that were compared to control LCLs obtained from a healthy donor (control 2, WT-LCL). We detected a substantial reduction in TRMT1 protein in the patient cell line from Family 16 containing a homozygous splicing variant predicted to abolish the splice acceptor site of exon 4 (Figure 4D, quantified in 4E). The patient from Family 19 harbors compound heterozygous missense variants *in trans* in *TRMT1*. We detected no substantial change in TRMT1 protein levels in the patient 19 LCL compared to control LCLs (Figure 4D, quantified in Figure 4E). These results suggest that the Arg323Cys (R323C) and Ser467Leu (S467L) missense variants do not significantly affect TRMT1 protein levels. Altogether, these results demonstrate that *TRMT1* splice variants as well as certain missense variants can impact TRMT1 protein accumulation.

### TRMT1-ID patient cells exhibit a reduction in m2,2G modification in tRNAs

We next tested the functional impact of *TRMT1* variants on tRNA modification in patient cell lines. TRMT1 has been shown to generate the m2,2G modification at position 26 in human tRNAs (6, 7). To monitor the m2,2G modification, we used a primer extension assay in which the presence of m2,2G leads to a block of reverse transcriptase (RT). A decrease in m2,2G modification allows for read-through and extension up to a subsequent RT-blocking modification. We performed the primer extension assay on tRNA-Met-CAU and mitochondria (mt)-tRNA-Ile-GAU, both of which contain m2,2G at position 26 (6, 17).

As reference, we performed the primer extension assay with RNA extracted from 293T human embryonic cells. In the absence of RT, only background bands were detected in reactions containing the radiolabeled probe and RNA from 293T human cells (representative gel shown in Figure 5A and B, lane 1). Addition of RT led to the appearance of an extension product up to the m2,2G modification at the expected position in both tRNA-Met-CAU and mt-tRNA-Ile-GAU in 293T human embryonic cells and a control fibroblast cell line from a healthy control with wildtype *TRMT1* alleles (Figure 5A and B, lanes 2 and 3, 293T and WT control). Patient fibroblast cells from Family 14 that are heterozygous for the D231N variant exhibit similar levels of m2,2G modification in tRNA-Met-CAU and mt-tRNA-Ile-GAU compared to the control fibroblast cell line (Figure 5A and B, compare lanes 3 and 4, quantified in Figure 5C and D). In contrast, patient fibroblast cells from Family 14 that are homozygous for the D231N variant exhibited nearly complete loss of the m2,2G modification block in tRNA-Met-CAU and mt-tRNA-Ile-GAU (Figure 5A and B, compare lane 5 to lanes 3 and 4; quantified in Figure 5C and D). These results indicate that the D231N variant impairs the methyltransferase activity of TRMT1 to form m2,2G.

**Figure 5.**
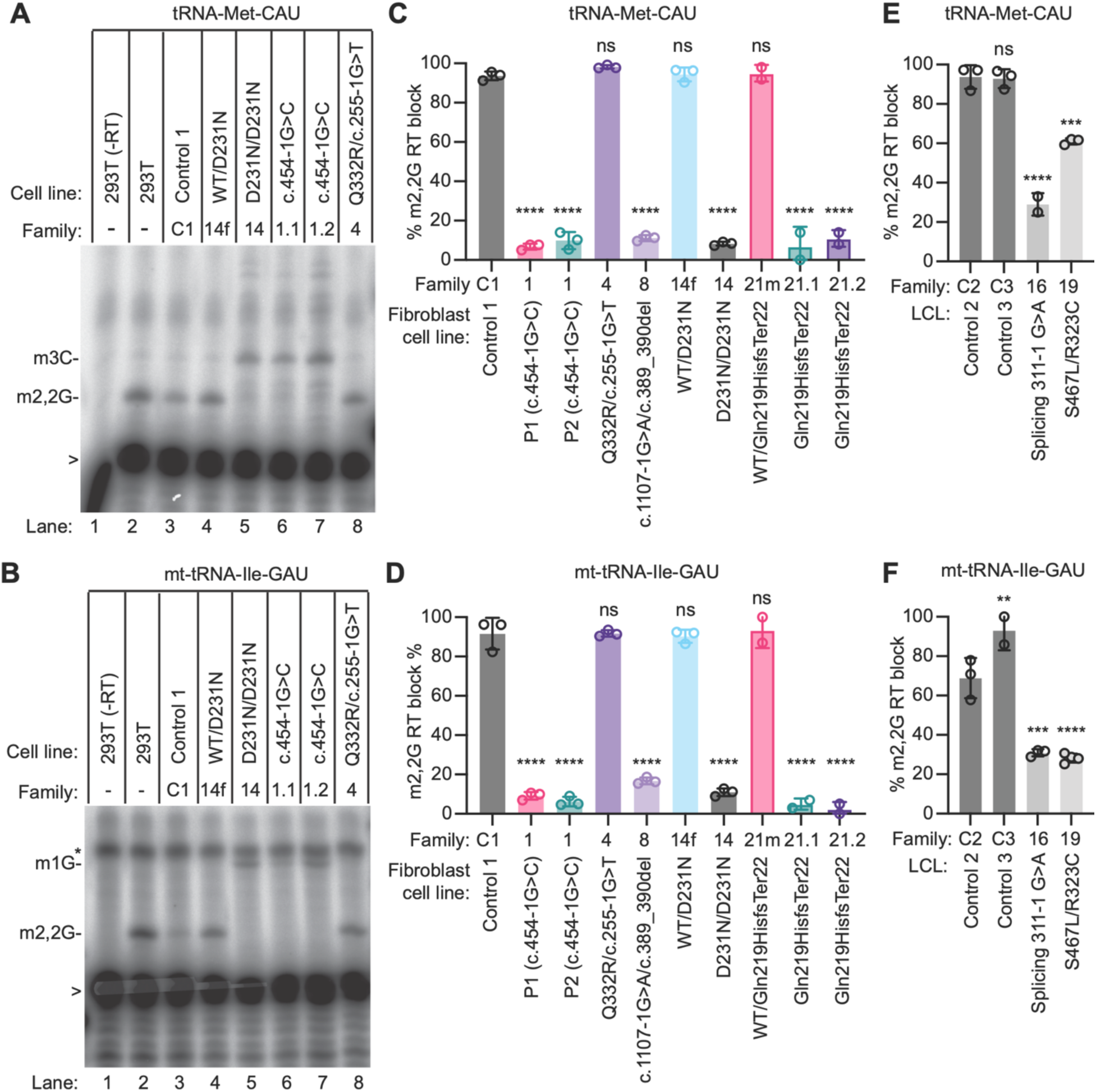
Patient cells with biallelic *TRMT1* variants exhibit a reduction in m2,2G modifications in tRNAs. (**A, B**) Representative gels of primer extension assays to monitor the presence of m2,2G in tRNA-Met-CAU and mt-tRNA-Ile-GAU from the indicated cell lines. m3C_20_, 3-methylcytosine; m2,2G_26_, dimethylguanosine; m1G_9_, 1-methylguanosine; >, labeled oligonucleotide used for primer extension; *, background signal. (**C through F**) Quantification of m2,2G formation by primer extension for the indicated tRNAs. N=3. Error bars represent standard deviation from the mean. Statistical analysis was performed using one-way ANOVA. For (C) and (D), the means of each column was compared to the Control 1 cell line. For (E) and (F), the means of each column was compared to the Control 2 cell line. *P ≤ 0.05; **P ≤ 0.01; ***P ≤ 0.001; ****P < 0.0001; ns, non-significant, P > 0.05.

The m2,2G modification in tRNA-Met-CAU and mt-tRNA-Ile-GAU was also reduced in patient-derived cell lines from families 1, 8, 16, 19, and 21 (Figure 5A and B, Supplementary Figure 3, quantified in Figure 5C through F,). The cell lines from families 1 and 16 are homozygous for *TRMT1* splicing variants, while the cell line from Family 8 is compound heterozygous for a splice site and frameshift variant. The cell lines from family 21 are derived from the mother (21m) and the mother’s children (21.1 and 21.2) who are heterozygous or homozygous for a *TRMT1* frameshift variant, respectively. The reduction in m2,2G modification in cell lines with homozygous splice site and/or frameshift variants is consistent with the loss of full-length TRMT1 protein expression. The patient cell line from Family 19 is compound heterozygous for the S467L/R323C missense variants. The reduction in m2,2G modification in this cell line indicates that the S467L and R323C variants reduce the activity of TRMT1.

No significant change in m2,2G modification was detected in the patient 4 cell line, which is compound heterozygous for the Q332R missense and a splice site variant (Figure 5A and B, lane 8, quantified in Figure 5C and D). These results suggest that the combination of these two *TRMT1* alleles produces enough active protein to maintain m2,2G modification in tRNA-Met-CAU and mt-tRNA-Ile-GAU. This finding is consistent with our observation that the patient 4 cell line exhibits comparable levels of TRMT1 protein as wildtype human cells (Figure 4).

### TRMT1 protein variants exhibit defects in reconstituting m2,2G modification in cells

We next used a TRMT1-knock out (KO) cell line derived from 293T human embryonic kidney cells to test TRMT1 variants for their ability to rescue m2,2G formation *in vivo*. The TRMT1-KO line lacks TRMT1 protein expression resulting in the absence of m2,2G modifications in all tested tRNAs (6). The TRMT1-deficient 293T cell line allowed us to further characterize the functionality of TRMT1-ID variants, including variants for which patient cell lines were not available. As a comparison, we also tested a TRMT1-S363L missense variant present as a minor allele in certain populations and is predicted to be non-pathogenic based upon mutation screenings (18).

Using transient transfection of plasmid constructs, we expressed either WT-TRMT1 or TRMT1 variants in the TRMT1-KO cell line. We then assessed for rescue of m2,2G formation in tRNA-Met-CAU or mt-tRNA-Ile-GAU using the primer extension assay described above. As expected, wildtype 293T cells transfected with vector alone exhibited an RT block at position 26 of tRNA-Met-CAU and mt-tRNA-Ile-GAU indicative of the m2,2G modification (Figure 6A through D, lane 1). The m2,2G modification was absent in tRNA-Met-CAU and mt-tRNA-Ile-GAU from the vector-transfected TRMT1-KO cell line leading to read-through to the next RT block (Figure 6A through D, lane 2). Re-expression of wildtype (WT)-TRMT1 in the TRMT1-KO cell line was able to restore m2,2G formation (Figure 6A through D, lane 3). Due to incomplete transfection efficiency which caused variable TRMT1 expression, the level of m2,2G modification was increased in the TRMT1-KO cell line but not completely rescued to the level of the original WT cell line.

**Figure 6.**
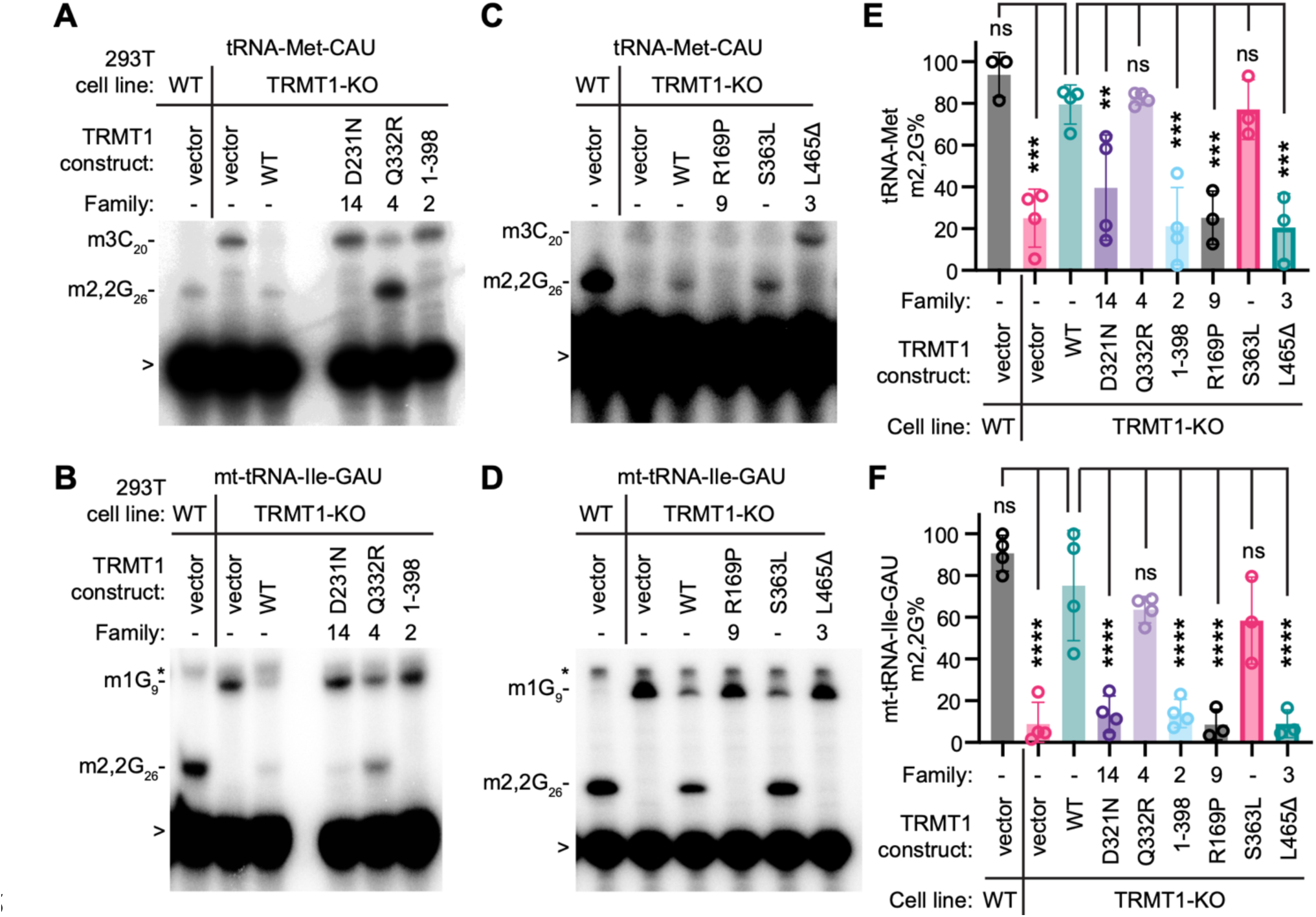
TRMT1 variants exhibit defects in reconstitution of tRNA modification activity in human cells. (**A through D**) Representative primer extension gels to monitor the presence of m2,2G in tRNA-Met-CAU and mt-tRNA-Ile-GAU from 293T cell lines transfected with the indicated constructs. m3C_20_, 3-methylcytosine; m2,2G_26_, dimethylguanosine; m1G_9_, 1-methylguanosine; >, labeled oligonucleotide used for primer extension; *, background signal. (**E, F**) Quantification of m2,2G formation by primer extension for the indicated tRNAs. Primer extensions were performed three times and error bars represent the standard error of the mean Statistical analysis was performed using one-way ANOVA. *P ≤ 0.05; **P ≤ 0.01; ***P ≤ 0.001; ****P < 0.0001; ns, non-significant, P > 0.05.

Using this assay, we found that the TRMT1-Q332R missense variant from Family 4 exhibited similar reconstitution of m2,2G formation as wildtype TRMT1 (Figure 6A and B, lane 5, quantified in Figure 6E and F). The wildtype activity of the Q332R variant is consistent with the wildtype levels of m2,2G modification detected in the tRNAs of the patient cell line derived from Family 4 (Figure 6A-D). The TRMT1-S363L minor variant also retained the ability to reconstitute m2,2G formation similar to that observed in WT-TRMT1 (Figure 6C and D, lane 5, quantified in Figure 6E and F).

Notably, we found that the TRMT1-D231N variant from Family 14 and TRMT1 truncation variant (1-398) from Family 2 were greatly reduced in their ability to reconstitute m2,2G formation in the TRMT1-KO cell line (Figure 6A and 6B, lanes 4 and 6, quantified in Figure 6E and F). The reduced activity of the TRMT1-D231N variant from Family 14 is consistent with the drastically reduced m2,2G levels in patient cells homozygous for the D231N variant (Figure 5). We also found that the R169P missense variant from Family 9 and TRMT1 L465 deletion variant from Family 3 exhibited defects in reconstituting m2,2G formation in TRMT1-KO cell lines (Figure 6C and 6D, lanes 4 and 6, quantified in Figure 6E and F). These results suggest that individuals homozygous for these variants are likely to be deficient in m2,2G modifications.

### TRMT1 variants exhibit defects in tRNA binding

We next investigated the interaction between TRMT1 variants and tRNAs to dissect the molecular defects associated with individual TRMT1-ID variants. We have previously shown that human TRMT1 displays a stable interaction with substrate tRNAs that are targets for m2,2G modification (6, 17). Using this system, we expressed a FLAG-tagged version of the TRMT1 variants in 293T human embryonic kidney cells followed by affinity purification and analysis of copurifying RNAs. Sample recovery of copurifying RNAs was confirmed through the spike-in addition of a synthetic RNA that served as a recovery control (RC). We analyzed the same set of TRMT1 variants as in Figure 6. Immunoblotting confirmed the expression and purification of each TRMT1 variant on anti-FLAG resin (Figure 7A, B).

**Figure 7.**
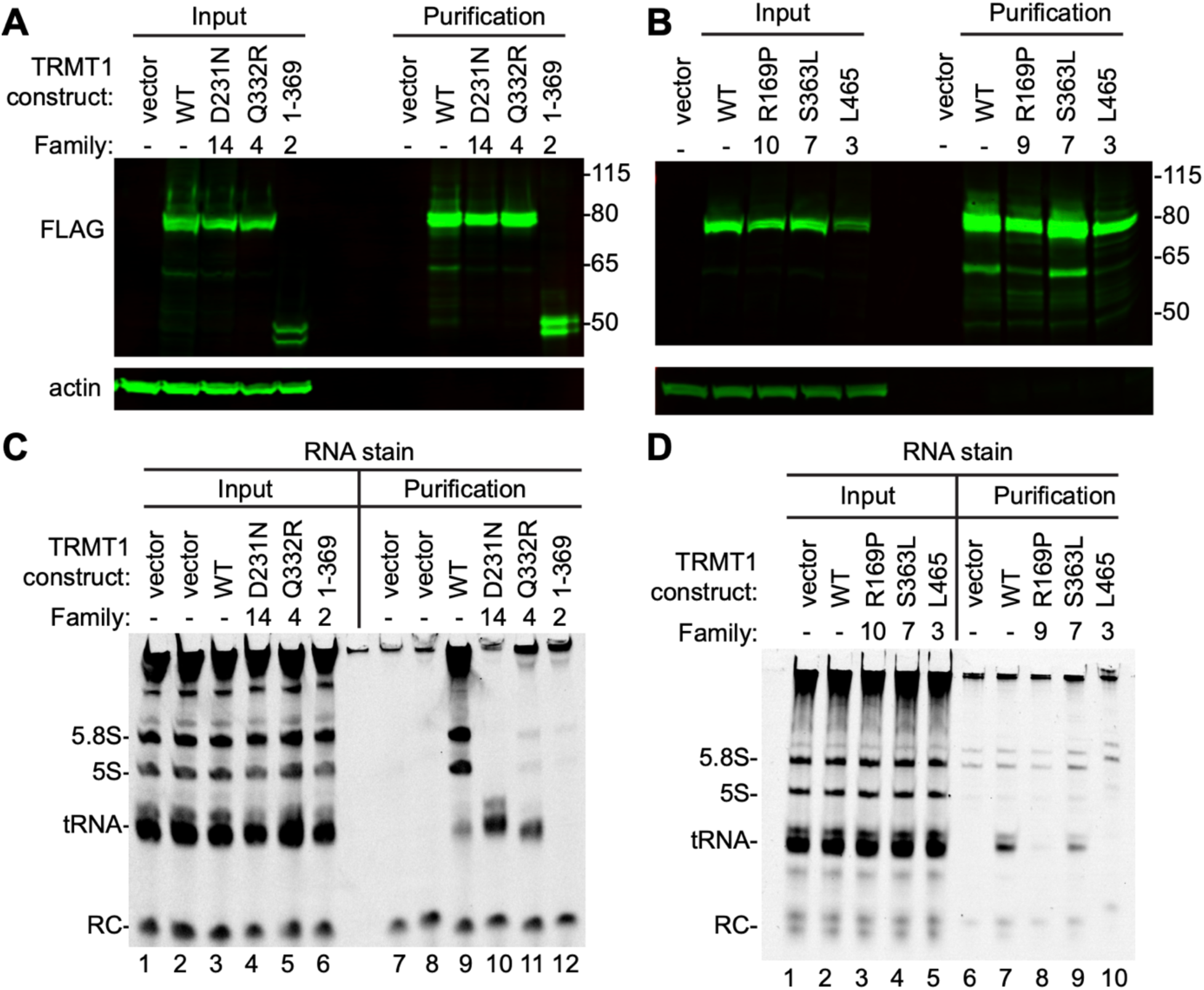
TRMT1-ID variants perturb the binding of TRMT1 to RNA. (**A, B**) Immunoblot of whole-cell extracts prepared from 293T human embryonic kidney cells transfected with each of the indicated FLAG-tagged constructs. Molecular weight in kilodalton is denoted on the right. (**C, D**) Nucleic acid stain of RNAs extracted from the indicated input or purified samples after denaturing polyacrylamide gel electrophoresis. The migration pattern of tRNAs, 5.8S, and 5S ribosomal RNA is denoted.

In the control purification from vector-transfected cells, we detected only background contaminating 5.8S and 5S ribosomal RNAs (5.8S and 5S; Figure 7C, lanes 7, 8; Figure 7D, lane 6). In contrast, the purification of WT-TRMT1 resulted in the enrichment of tRNAs along with rRNAs as we have previously shown (Figure 7C, lane 9; Figure 7D, lane 7). In contrast to WT-TRMT1, we found that the TRMT1 truncation variant found in Family 2 exhibited defects in binding RNA (Figure 7B, lane 12). Interestingly, we found that the TRMT1-D231N and Q332R variants exhibited increased copurification of tRNAs compared to wildtype TRMT1 while rRNA copurification was decreased (Figure 7C, compare lane 9 to lanes 10 and 11). These results suggest that the D231N and Q332R variants alter the RNA binding specificity of TRMT1 to prefer interaction with tRNAs versus rRNAs.

We found that the TRMT1-R169P missense variant from Family 10 and L465 deletion variant found in Family 3 exhibited reduced binding to tRNAs compared to wildtype TRMT1 (Figure 7D, lanes 8 and 10). The reduced tRNA binding by the TRMT1-R169P could explain its diminished ability to reconstitute m2,2G formation in cells. The TRMT1-S363L minor variant exhibited similar binding to tRNAs compared to wildtype TRMT1. The wildtype tRNA binding of the TRMT1-S363L variant is consistent with the wildtype activity of this variant in reconstitution assays observed above. Altogether, these findings uncover the molecular effects of ID-associated TRMT1 variants on methyltransferase activity and tRNA binding that underlie deficits in m2,2G modification in patient cells.

### TRMT1 missense variants reveal distinct functional regions required for TRMT1 activity

To gain insight into the functional effects, we mapped the *TRMT1* variants onto a predicted human TRMT1 structure generated through AlphaFold (19). The hypothesized structure of human TRMT1 was aligned with the solved structure of Trm1 bound to SAM from the archaea *Pyrococcus horshiki* (20). Based upon this structural alignment, human TRMT1 is predicted to fold into two domains coinciding with the SAM-dependent methyltransferase domain and a C-terminal domain unique to Trm1 enzymes (Figure 8, N-terminal domain in blue, C-terminal domain in yellow). The N-terminal domain of TRMT1 forms a putative active site for binding of the SAM methyl donor and a pocket for accommodating the G26 nucleotide that undergoes methylation (Figure 8, red dash circle denotes active site, SAM denoted in green).

**Figure 8.**
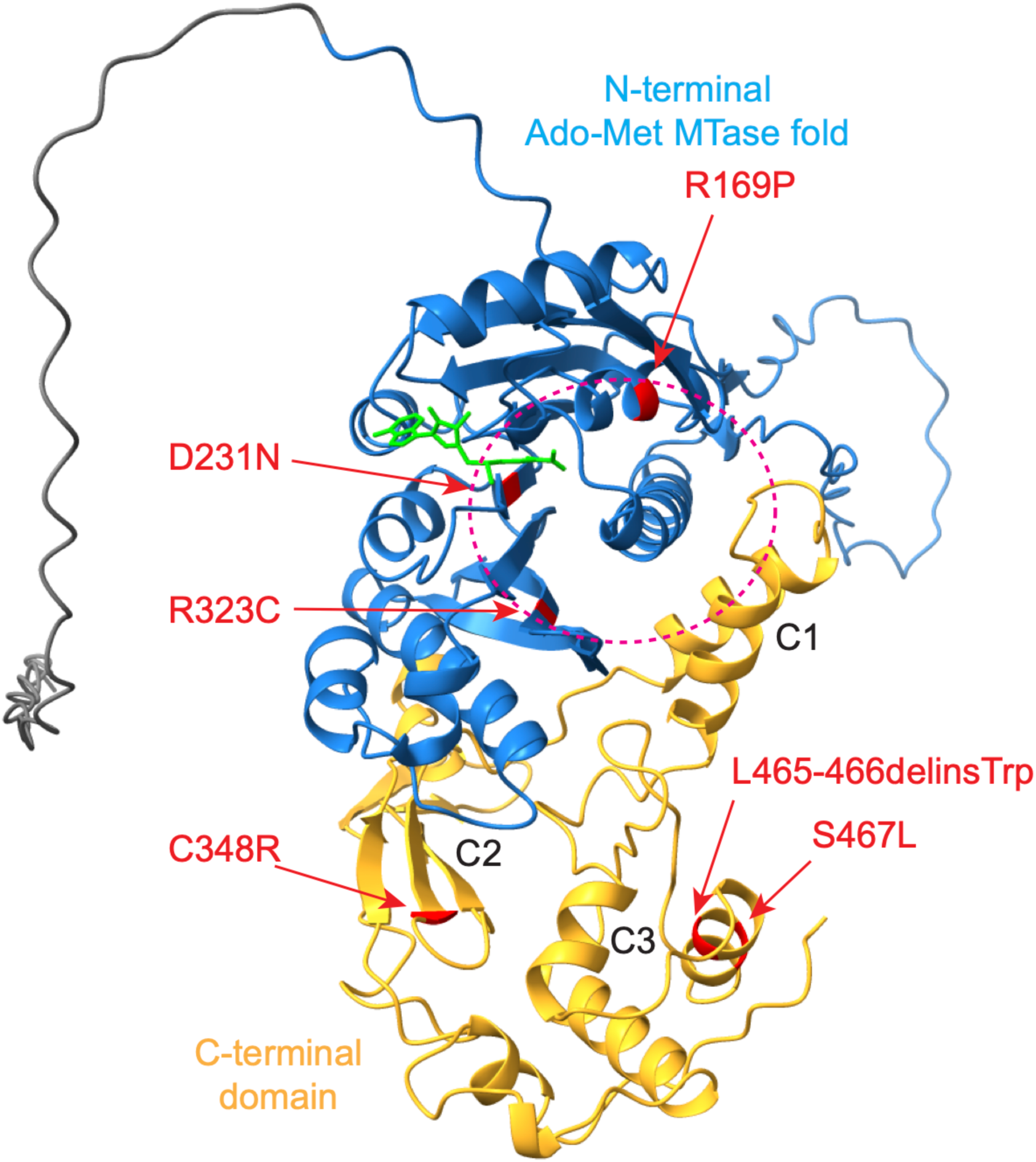
Location of missense variants in the predicted structure of human TRMT1. The methyltransferase domain along with the unstructured N-terminus (gray) are shown while the C-terminal zinc finger motif has been omitted. The human TRMT1 model was aligned with *Pyrococcus horikoshii* Trm1 bound to *S*-adenosyl-methionine (PDB: 2EJT) and domains are colored according to (20). The N-terminal Adenosyl-methionine-dependent methyltransferase domain is depicted in blue and C-terminal domain in yellow. Dashed circle represents the putative catalytic active site for binding and methylation of the G26 nucleotide in substrate tRNAs. *S*-adenosyl-methionine is denoted in green. The locations of TRMT1 variants are noted in red.

Notably, the D231N and R323C variants are situated near the predicted G26 pocket (Figure 8, D231N and R323C). As shown above, the D231N and R323C variants are defective in tRNA modification activity but retain similar levels of tRNA binding as wildtype TRMT1. This result is consistent with these variants perturbing G26 substrate positioning in the active site and preventing catalysis without a major effect on overall tRNA recognition and binding. Similar to the R323C and D231N variants, the R169P variant lies nearby the putative G26 binding pocket of TRMT1. However, in contrast to the R323C and D231N variants, the R169P variant is predicted to disrupt the formation of a conserved alpha helix within the active site that is likely to cause broader changes in the N-terminal domain. This drastic alteration in structure is consistent with the R169P variant exhibiting defects in both tRNA modification activity and tRNA binding (Figure 7).

The C348R, L465-R466deletion, and S467L variants lie within the C-terminal domain that is unique to the Trm1 enzyme family. The C348R variant resides within the C1 subdomain. In *Pyrococcus horshiki* Trm1, the C1 subdomain makes numerous hydrophobic contacts with the N-terminal domain (20). Thus, the C348R variant could alter the folding of the C1 subdomain thereby impacting the N-terminal catalytic domain. The L465R and S467L variants lie within a predicted alpha helix of the C3 subdomain, which faces across from the active site (Figure 8, C3). The C3 subdomain exhibits similarity with subdomains in phenylalanine tRNA synthetase that bind the anticodon region of tRNA-Phe (21). This similarity suggests that the C3 subdomain of TRMT1 could form additional contacts with the tRNA anticodon domain during substrate binding. Consistent with this role, we have found that the L465R variant disrupts tRNA binding and reconstitution of tRNA modification activity. Altogether, the TRMT1 missense variants reveal distinct functional activities linked to specific subdomains within the TRMT1 polypeptide.

### Depletion of Trmt1 in zebrafish causes developmental and behavioral perturbations

To investigate the loss of TRMT1 function *in vivo*, we used zebrafish as a model and employed the CRISPR/Cas9 method to generate biallelic mutations using three guide RNAs targeting the functional domain. We analyzed the phenotype in the F0 (founder) generation because our previous data suggest that F0 knockouts recapitulate phenotypes from the stable genetic knockouts (22, 23). RT-qPCR results found significant downregulation of *trmt1* mRNA expression in F0 knockouts (Figure 9A). We then used liquid chromatography-mass spectrometry to measure m2,2G levels in whole larvae or head-only samples from Cas9-injected control and *trmt1* F0 knockout larvae. Consistent with the depletion of Trmt1, the levels of m2,2G modification were reduced in both whole larvae or head-only samples from *trmt1* F0 knockout larvae (Figure 9B). In contrast, no significant difference was detected in the levels of the 1-methyladenosine (m1A) modification, which is another widespread tRNA modification (Figure 9C). We performed morphological phenotyping and found that *trmt1* F0 fish exhibited reduced head, eye, and body sizes (Figure 9D to G). These results indicate that depletion of Trmt1 and reduction in m2,2G modifications is sufficient to induce a general developmental delay in zebrafish.

**Figure 9.**
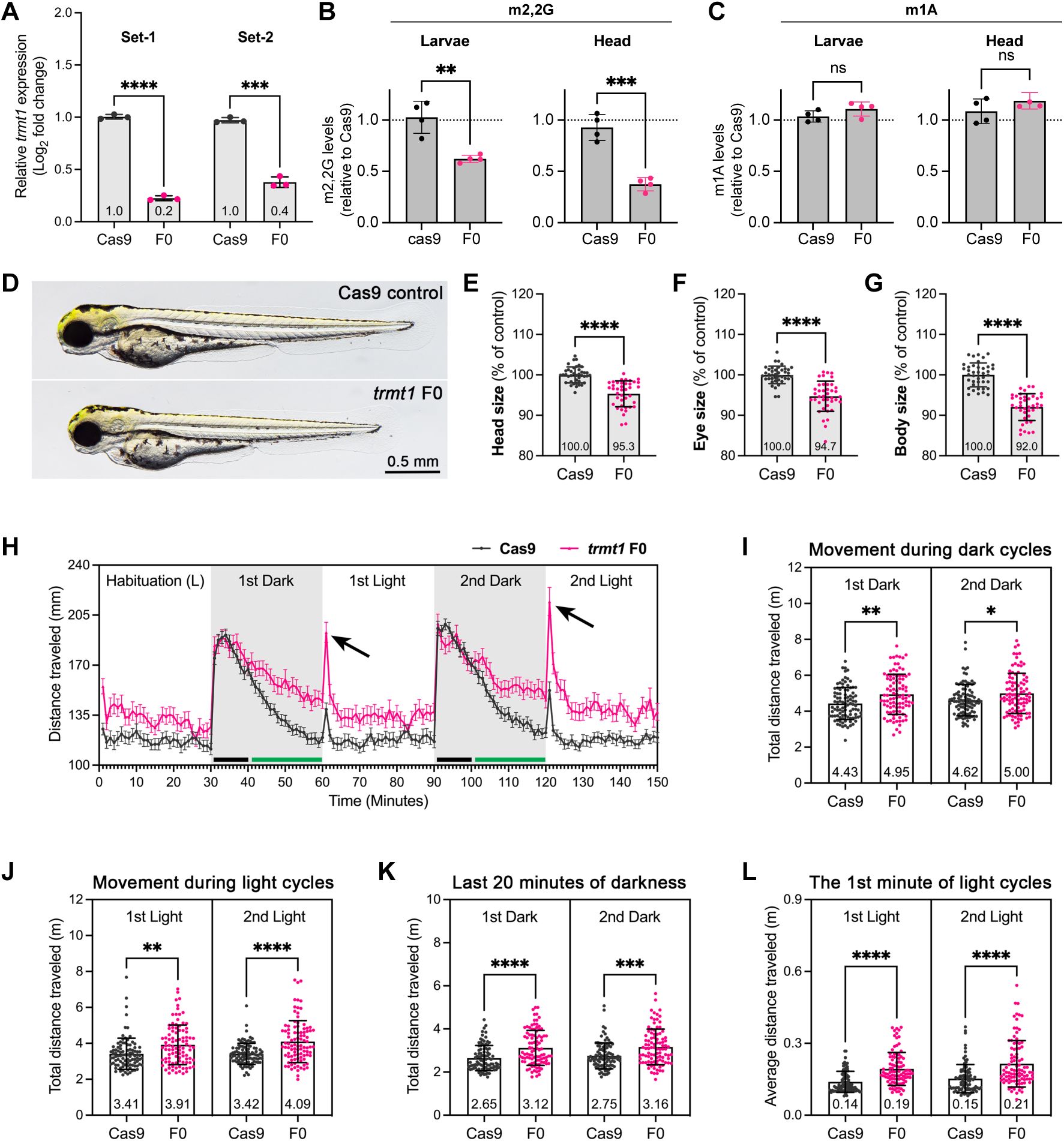
Depletion of Trmt1 in zebrafish induces developmental delay and behavioral abnormalities. (**A**) RT-qPCR was performed using two sets of primer pairs to examine the expression levels of *trmt1* mRNA in Cas9 control and *trmt1* F0 knockout animals at 4 dpf. Experiments were performed with biological and technical triplicates. Expression levels were normalized to the *18S* housekeeping gene and compared to the Cas9 animals. Error bar = mean ± SD. (**B, C**) LC-MS analysis of m2,2G or m1A in whole larvae or head-only samples. (**D**) Representative images for Cas9-injected control (Cas9) and *trmt1* F0 knockouts (F0) at 3 dpf. (**E-G**) Quantifications of head, eye, and body sizes for Cas9 and *trmt1* F0 animals. n = 40 embryos each. The values are presented as a percentage of the mean value of Cas9 controls. Error bars = mean ± SD. Each dot represents one larva. (**H**) Locomotor activity analysis, 96-well plates containing 96 larvae at 5 dpf were placed in a recording chamber. Larvae were habituated in the light for 30 minutes, followed by two 30-minute cycles of alternating dark and light cycles. Each point represents the average distance traveled by the animals, with n = 96 larvae each. Error bars indicate mean ± SEM. Black arrows indicate the first minute of light cycles. Black bars at the bottom indicate the first 10 minutes and green bars indicate the last 20 minutes of dark cycles. (**I**) Total distance traveled of each larva in the dark cycles. (**J**) Total distance traveled of each larva in the light cycles. (**K**) Total distance traveled of each larva in the last 20 minutes of dark cycles. (**L**) Average distance traveled calculated for each larva in the first minute of dark cycles. Error bars indicate mean ± SD. Each dot represents one larva. Mean value of each quantification was presented at the bottom of bar. For (B-D, F-I), statistical significance was calculated by unpaired *t* test with Welch’s correction: \**p* < 0.05, \*\**p* < 0.01, \*\*\**p* < 0.001 and \*\*\*\**p* < 0.0001.

Given that many affected individuals with pathogenic *TRMT1* variants exhibit behavioral phenotypes, zebrafish larvae were used to identify distinct swimming patterns in response to light and dark conditions. Therefore, we conducted a behavioral assay by exposing zebrafish larvae to alternating 30-minute light and dark cycles. Interestingly, we found that the mutant larvae displayed increased overall locomotor activity in both light and dark cycles (Figure 9H to J). During the dark cycles, the F0 knockouts showed similar activity to control fish in the initial 10 minutes (black bars in Figure 9H, quantified in Supplementary Figure 4A), but sustained higher activity in the remaining 20 minutes (green bars in Figure 9H, quantified in Figure 9K), suggesting a hyperactivity-like behavior. Additionally, the mutants showed a pronounced increase in movement during the first minute of light cycles (Figure 9L), potentially indicating light-induced seizure-like behavior (22). Furthermore, a decreased acoustic-evoked behavioral response (AEBR) was observed in the *trmt1* knockout larvae (Supplementary Figure 4B), suggesting impaired hearing. The phenotype analysis in F0 knockouts recapitulates a subset of patient symptoms, demonstrating a conserved role for Trmt1 in zebrafish.

## Discussion

In this study, we identify and characterize novel variants in *TRMT1* that impact mRNA splicing, protein levels and/or enzymatic activity. Our studies define a core set of phenotypic features universally associated with pathogenic *TRMT1* variants that encompasses intellectual disability, global developmental delay, and facial dysmorphism. While no major intrafamilial phenotypic variability was observed, the present cohort exhibited remarkable interfamilial phenotypic variability characterized by a spectrum of behavioral, morphological, and physiological features. These findings are significant by indicating that TRMT1 is required for a common set of developmental and neurological pathways with further clinical outcomes determined by genetic and environmental factors specific to each family.

Depletion of Trmt1 and m2,2G modifications in zebrafish results in developmental and behavior phenotypes that resemble certain core features of TRMT1-associated syndromes in human patients. Importantly, these findings demonstrate that TRMT1 homologs play a conserved role in the proper development and function of the central nervous system in vertebrates. The future generation of zebrafish strains that are biallelic for the pathogenic variants discovered here will allow us to further dissect the spectrum of phenotypes in an isogenic background.

Overall, our findings support a model in which loss of m2,2G modifications due to a decrease in functional TRMT1 protein and/or activity results in downstream perturbations in molecular and cellular processes that cause neurodevelopmental phenotypes. Moreover, we find that the penetrance of the core phenotypic effects can depend on the severity of the variant on TRMT1 function in tRNA modification as well as the specific type of change caused by the TRMT1 variant. For example, we find that TRMT1 variants can induce aberrant splicing, but with distinct outcomes that could differentially impact the functional levels of TRMT1. In addition to loss-of-function splice isoforms, there could be aberrant splice variants that exert dominant negative effects by coding for TRMT1 products that bind tRNA substates without modifying them or exhibit gain-of-function properties. It will also be interesting to determine if any of the splice variants serve regulatory roles that are perturbed by the *TRMT1* variants.

Since TRMT1 is known to modify numerous tRNA targets, each tRNA could be affected to a different extent by a *TRMT1* variant that could account for the variable phenotypic outcomes. For example, the compound heterozygous S467L/R323C variant appears to more severely impact the modification of mt-tRNA-Ile compared to cytoplasmic tRNA-Met. In addition, the TRMT1 protein variants that affect catalytic activity without impacting tRNA binding could retain RNA chaperone functions that are completely abrogated for other TRMT1 protein variants. Future studies that profile the global modification status and levels of individual tRNAs in each patient would shed light on the differential effects of each TRMT1 variant.

The variable clinical presentations and age of onset of individuals with similar genotypes support the existence of additional, currently unidentified modifying variants in other genes besides TRMT1. Future studies will focus on identifying genetic modifiers in this cohort that could reveal the biological pathways and processes that are connected to TRMT1 function. Importantly, the functional demonstration of pathogenicity for so many variants in multiple families across the world indicates that *TRMT1* should be included in genetic registries as a key disease gene linked to developmental brain disorders with autosomal recessive Mendelian inheritance (24).

## MATERIAL AND METHODS

### Recruitment of research subjects

The 24 families with bi-allelic *TRMT1* variants were identified using the GeneMatcher platform (25) and data sharing with collaborators. Informed consent for genetic analyses was obtained from all subjects. Clinical details of the cohort were obtained by the follow-up of affected individuals. Seizure description is reported in line with the most recent ILAE guidance (26). Parents and legal guardians of all affected individuals gave their consent for the publication of clinical and genetic information according to the Declaration of Helsinki, and the study was approved by The Research Ethics Committee Institute of Neurology University College London (IoN UCL) (07/Q0512/26) and the local Ethics Committees of each participating center. Consent has been obtained from a subset of families to publish medical photographs and video examinations. Brain magnetic resonance imaging (MRI) scans were obtained from 12 affected individuals and were reviewed by an experienced team of pediatric neuroradiologists. Cerebellar atrophy, callosal thinning, and calvarial deformities were defined using standardised criteria (27–29). Facial photographs and/or videos of 13 patients from 10 families were reviewed and their dysmorphic features were described using terminology recommended by Elements of Morphology. Where no term was available for a dysmorphic feature seen in a patient, HPO terminology was used instead.

### Variant identification and NGS data interpretation

Single-nucleotide variations (SNVs) were identified by Whole Exome Sequencing (WES) or Whole Genome Sequencing (WGS) in all individuals. Exomes or genomes were captured and sequenced on Illumina sequencers as described elsewhere (30) in Macrogen, Korea or at collaborating centers (see Table 2). The bioinformatics filtering strategy included screening for only exonic and donor/acceptor splicing variants. Rare variations present at a frequency above 1% in GnomAD v.3.1.2 (https://gnomad.broadinstitute.org/) or present from exomes or genomes within datasets from UK Biobank and UK 100,000 genome project or from internal research databases (*e.g.*, Queen Square Genomics and UCL SYNaPS Study Group) were excluded. Candidate variants were then inspected with the Integrative Genomics Viewer and then confirmed by Sanger sequencing in all the families. Sequence variants in the *TRMT1* gene were described according to the recommendations of HGVS and are based on the reference sequence NM_001136035. Sequence candidate variants were interpreted according to ACMG Guidelines (31).

### Cell culture of primary dermal fibroblasts

Primary dermal fibroblasts were obtained from a skin biopsy of subjects. Fibroblasts were cultured in Dulbecco’s modified Eagle medium (DMEM; Thermo Fisher Scientific) supplemented with 10% fetal bovine serum (FBS; GE Healthcare) and penicillin-streptomycin (100 U/mL and 100 mg/mL, respectively; Thermo Fisher Scientific). For all experiments, the same passage number of subject and control fibroblasts was used. Primary fibroblasts were regularly tested for mycoplasma contamination and confirmed to be mycoplasma free.

### Cell culture of primary lymphoblasts

Lymphoblastoid cell lines (LCLs) are generated by Epstein–Barr virus (EBV) transformation of the B-lymphocytes within the peripheral blood lymphocyte (PBL) of patients. LCLs were cultured in cells in RPMI 1640 medium (Thermo Fisher Scientific) supplemented with 10% fetal bovine serum (FBS; GE Healthcare) and penicillin-streptomycin (100 U/mL and 100 mg/mL, respectively; Thermo Fisher Scientific) in standing flasks at 37°C and 5% CO2 for several days until adhering to the flask and reaching a desired cell count.

### Minigene splicing assay

Computational assessment of splicing effects used SpliceSiteFinder-like, MaxEntScan, NNSplice, and GeneSplicer embedded in Alamut Visual Plus v1.6.1 (Sophia Genetics, Bidart, France), as well as SpliceAI 10K and AbSplice as included in SpliceAI Visual (32).

RNA studies of variants were conducted following established protocols with some modifications (33, 34) using three constructs with variants annotated to NM_001136035.4. In brief, the first construct comprised a 1002 bp region spanning introns 2 to 5, encompassing the c.255-1G>T, c.310+5G>C, c.311-1G>A, and c.454-1G>C variants. The second construct involved a 416 bp segment spanning introns 8 to 10 to assay splice effects of the c.1107-1G>A variant. Finally, the third construct covered a 446 bp region spanning intron 10 to exon 12, targeting the c.1194G>A variant. These regions were amplified from genomic DNA obtained from the probands and a healthy control using primers containing specific restriction sites (Supplementary Table 7). The PCR fragments were ligated between exons A and B of the linearized pSPL3-vector following digestion with restriction enzymes. The recombinant vectors were transformed into DH5α competent cells (NEB 5-alpha, New England Biolabs, Frankfurt, Germany), plated and incubated overnight. Following colony PCR with SD6 F (Supplementary Table 7) and the target-specific reverse primer, the wild-type and mutant-containing vector sequences were confirmed by Sanger sequencing and transfected into HEK 293T cells (ATCC, Manassas, VA, USA). 2 µg of the respective pSPL3 vectors was transiently transfected using 6 µL of FuGENE 6 Transfection Reagent (Promega, Walldorf, Germany). An empty vector and transfection negative reactions were included as controls. The transfected cells were harvested 24 hours after transfection. Total RNA was isolated using miRNeasy Mini Kit (Qiagen, Hilden, Germany). cDNA was synthesized using the High Capacity cDNA Reverse Transcription Kit (Applied Biosystems, Waltham, MA, USA) following the manufacturer’s protocols. cDNA was PCR amplified using vector-specific SD6 F and SA2 R primers (Supplementary Table 7). The amplified fragments were visualized on a 1% agarose gel. cDNA amplicons were TA cloned following standard protocols with the pCR2.1 vector kit (ThermoFisher, Darmstadt, Germany) and Sanger sequenced. Fragment analysis was performed for construct 1 with FAM-labelled SD6 F and SA2 R primers using the 3500xL Genetic Analyzer (Thermo Fisher Scientific, Waltham, MA, USA). Analysis was performed using GeneMapper Software 5 (Applied Biosystems). Analysis and cataloging of protein-coding versus non-protein coding transcripts and their expression was performed using Ensembl and GTEx Portal. Non-coding transcripts were excluded in calculations to determine the average peak area for construct 1. The percentage of each band was calculated using fragment analysis (Supplementary Figure 2, Supplementary Data 1).

### Immunoblotting

For protein immunoblotting, fibroblast or lymphoblast cells were resuspended in hypotonic lysis buffer for protein extraction as noted previously (17, 35, 36). Cell extracts were boiled at 95°C for 5 minutes followed by fractionation on NuPAGE Bis-Tris polyacrylamide gels (Thermo Scientific). Separated proteins were transferred to Immobilon FL polyvinylidene difluoride (PVDF) membrane (Millipore) for immunoblotting. Membrane was blocked by Odyssey blocking buffer for 1 hour at room temperature followed by immunoblotting with the following antibodies: anti-TRMT1 (sc-373687, Santa Cruz Biotechnology), anti-FLAG epitope tag (L00018; Sigma) and anti-actin (L00003; EMD Millipore). Proteins were detected using a 1:10,000 dilution of fluorescent IRDye 800CW goat anti-mouse IgG (925-32210; Thermofisher).

### RNA analysis

RNA was extracted using TRIzol LS reagent (Invitrogen). For primer extension analysis, 1.5 μg of total RNA was pre-annealed with 5’-32P-labeled oligonucleotide and 5x hybridization buffer (250 mM Tris, pH 8.5, and 300 mM NaCl) in a total volume of 7 μl. The mixture was heated at 95°C for 3 min followed by slow cooling to 42°C. An equal amount of extension mix consisting of avian myeloblastosis virus reverse transcriptase (Promega), 5x AMV buffer and 40 μM dNTPs was added. The mixture was then incubated at 42°C for 1 hour and loaded on 18 to 20% 7M urea denaturing polyacrylamide gels. Gels were exposed on a phosphor screen and scanned on a Sapphire Biomolecular Imager (Azure Biosystems). Quantification was performed using NIH ImageJ software followed by statistical analysis using GraphPad Prism. Primer extension oligonucleotide sequences were previously described (Dewe et al., 2017).

### Transient transfection of 293T cells

The 293T TRMT1-KO cell line has been described previously (6). 293T cells were transfected via calcium phosphate transfection method (37). Briefly, 2.5 × 10^6^ cells were seeded on 100 × 20 mm tissue culture grade plates (Corning) followed by transfection with 10 μg of plasmid DNA. Cells were harvested 48 h later by trypsin and neutralization with media, followed by centrifugation of the cells at 700 × g for 5 min followed by subsequent PBS wash and a second centrifugation step.

### Protein-RNA purifications

Protein was extracted using hypotonic lysis and high salt immediately after cells were harvested. Cell pellets were resuspended in 0.5 ml of hypotonic lysis buffer (20 mM HEPES pH 7.9, 2 mM MgCl2, 0.2 mM EGTA, 10% glycerol, 0.1 mM PMSF, 1 mM DTT) per 100 × 20 mm tissue culture plate. Cells were kept on ice for 5 min and then subjected to 3 freeze-thaw cycles in liquid nitrogen and a 37 °C water bath. NaCl was then added to the extracts at a concentration of 0.4 M, incubated on ice for 5 mins, and centrifuged at 14,000 × g for 15 min at 4 °C. After centrifugation, 500 μl of supernatant extract was removed and 500 μl of Hypotonic Lysis buffer supplemented with 0.2% NP-40 was added to obtain 1,000 μl of extract.

FLAG-tagged proteins were purified by incubating whole cell lysates from the transiently-transfected cell lines with 50 μl of Anti-DYKDDDDK Magnetic Beads (Clontech) for two hours at 4 °C. Magnetic resin was washed three times in hypotonic wash buffer (20 mM HEPES pH 7.9, 2 mM MgCl2, 0.2 mM EGTA, 10% glycerol, 0.1% NP-40, 0.2 M NaCl, 0.1 mM PMSF, and 1 mM DTT). SDS-PAGE sample buffer was added to one portion of resin and purified proteins were fractionated on a NuPAGE Bis-Tris polyacrylamide gel (ThermoFisher). The gel was transferred to Immobilon-FL Hydrophobic PVDF Transfer Membrane (Millipore Sigma) with subsequent immunoblotting against the FLAG tag as noted above.

RNA from input and purified samples were extracted using RNA Clean & Concentrator-5 columns (Zymo Research). RNA extraction followed TRIzol LS RNA extraction protocol (Invitrogen). RNA was resuspended in 5 μl of RNAse-free water and loaded onto a 10% polyacrylamide, 7 M urea gel. The gel was then stained with SYBR Gold nucleic acid stain (Invitrogen) to visualize RNA.

### Ethics Statement and Zebrafish Husbandry

All experimental animal care was performed in accordance with institutional and NIH guidelines and regulations. Zebrafish (*Danio rerio*) were raised and maintained in an Association for Assessment and Accreditation of Laboratory Animal Care (AAALAC)-accredited facility at the Oklahoma Medical Research Foundation (OMRF) under standard conditions. All experiments were conducted as per protocol (22–76) approved by the Institutional Animal Care Committee (IACUC) of OMRF.

### Generation of *trmt1* F0 knockout zebrafish

We followed previously described methods to generate *trmt1* F0 knockouts (PMID: 38031187). Briefly, three guide sequences were designed using the CRISPOR tool, and guide RNAs (gRNAs) were chemically synthesized (Synthego Inc., CA, USA). A 6 μL mixture containing 1 μL of 40 μM Cas9-NLS protein (UC Berkeley QB3 Macrolab, Berkeley, CA, USA), 500 ng of each gRNA (in 3 μL), and 2 μL of 1 M potassium chloride was injected into one-cell stage wild-type (WT) embryos. As a control, WT embryos were injected with a mixture containing Cas9 protein but lacking gRNA. The gRNA target sequences are listed in Supplementary Table 8.

### RNA extraction and reverse transcription-quantitative PCR (RT-qPCR)

Total RNA was extracted from either whole larvae or head-only samples using TRIzol Reagent (Thermo Fisher Scientific, USA) and purified with the RNA Clean and Concentrator-5 kit (Zymo, USA) according to the manufacturer’s instructions. Larvae at 4 dpf were anesthetized in 168 mg/L Tricaine methanesulfonate/MS-222 (Sigma-Aldrich, MO, USA) before head dissection. Each experimental group consisted of three biological replicates, with six larvae pooled per replicate, collected randomly at 4 days post-fertilization (dpf). cDNA was synthesized using the iScript RT Supermix (Bio-Rad, CA, USA) and used as a template for RT-qPCR with SYBR Green Supermix (Thermo Fisher Scientific, CA, USA) on the Light Cycler® 96 System (Roche, CA, USA). All RT-qPCR reactions were performed in biological triplicates with technical triplicates. The *18S* gene was used as a reference gene. Primer sequences for RT-qPCR are listed in Supplementary Table 8. Cycle threshold (Ct) values were analyzed in Microsoft Excel, and relative gene expression was quantified using the 2^(-ΔΔCt) method.

### Morphological phenotyping

To assess morphological phenotypes, zebrafish larvae were randomly selected at 4 dpf for imaging. Larvae were manually positioned in 2% methylcellulose (Sigma, USA) under a stereomicroscope and imaged. Head, eye, and body measurements were obtained directly from scale-calibrated images using ImageJ software (NIH). Head size was determined by measuring the distance from the tip of the snout to the end of the operculum (gill cover). Eye size was measured as the eye diameter, and body size was measured as the length from the tip of the snout to the end of the tail. All bright-field images were captured using a Nikon DS-Fi2 high-definition camera mounted on a Nikon SMZ18 stereomicroscope (Nikon, Japan) with auto-Z-stacking capability.

### Behavioral assay

All behavior tests were performed at room temperature (RT), as previously described (22). In brief, to conduct the LDT test, larvae at 4 dpf were delicately moved into individual wells of a 96-well plate, each containing 150 µL of embryo water. The following day, the plate was placed into a Noldus chamber, and locomotion activity was recorded using the DanioVision system, which runs EthoVision XT software (Noldus Information Technology, Leesburg, VA, USA). Specifically, larvae at 5 dpf were allowed a 30-minute habituation period in the light, followed by alternating 30-minute dark and light periods for two cycles. The locomotion activity of the larvae was measured in terms of distance traveled (in millimeters) per minute. The recorded values for each minute were then plotted using GraphPad Prism (GraphPad Software, San Diego, CA, USA). Larvae at 6 dpf were subjected to an acoustic-evoked behavioral response (AEBR) test using the Zebrabox (ViewPoint Life Sciences, Montreal, Canada). For AEBR quantification, the number of responses to 12 stimuli for each larva was calculated as a percentage of the total responses. Values were plotted using box and whisker plot generated by GraphPad Prism. Error bars indicate the range from the minimum to the maximum values, with the median value represented by the line in the center of the box.

### Statistics

The statistical analysis was conducted using GraphPad Prism. Data are presented as indicated in figure legends. For all analyses, the significance level was set at 0.05. Significance was determined using a two-tailed unpaired Student’s *t*-test with Welch’s correction for two comparisons, as detailed in the figure legends. *P*-values were represented as follows: not significant (ns) *p* ≥ 0.05, \**p* < 0.05, \*\**p* < 0.01, \*\*\**p* < 0.001, and \*\*\*\**p* < 0.0001.

## Acknowledgements

The authors would like to thank the affected individuals and their families for their support of this study. One of the authors of this publication (ZT) is a member of the European Reference Network on Rare Congenital Malformations and Rare Intellectual Disability ERN-ITHACA [EU Framework Partnership Agreement ID: 3HP-HP-FPA ERN-01-2016/739516]. BV is a member of the European Reference Network on Rare Congenital Malformations and Rare Intellectual Disability (ERN-ITHACA) [EU Framework Partnership Agreement ID: 3HP-HP-FPA ERN-01-2016/739516].

## Data availability

The data that supports the findings of this study are available within the paper and in the supplementary material. Whole-exome sequencing data are not publicly available due to privacy or ethical restrictions. The novel *TRMT1* variants reported in this manuscript were submitted to the LOVD database (https://databases.lovd.nl/shared/genes/TRMT1), with the LOVD variant IDs: # 0000944528, # 0000944622, # 0000944624, # 0000944625, # 0000944626, # 0000944640, # 0000944641, # 0000944642, # 0000944643, # 0000944646, # 0000944620, # 0000944621, # 0000944647, # 0000944709, # 0000944708, # 0000944710, # 0000944712, # 0000944713, # 0000944714, # 0000944715, # 0000944716, #0000959740, #0000959741, #0000959742, #0000959743.

## Author contributions

Conceptualization: S.E., A.E.F., R.M., D.F.; Data Curation: S.E., C.D., C.L., K.Z., S.J.L, R.Q.L, I.K., D.F.; Formal Analysis: S.E., C.D., C.L., K.Z., R.Q.L, I.K. D.O., J.S., K.M. B.V., D.F.; Methodology: S.E., B.V., D.F.; Funding Acquisition: B.V., G.V., H.H., D.F.; Investigation: all authors; Recruitment, Clinical, and Diagnostic Evaluations: R.K., F.J., J.R.A., T.S., C.L., M.J., F.T., M.I.V.P., R.S., G.Y., M.O., J.F., E.H.G., C.P., B.I., C.P., S.H., D.S., V.B., K.P., D.W., M.K., N.R., A.T., H.M., C.F., S.T.B., A.B., N.C., G.L., S.C., Z.T., T.D.H., G.R., T.M., J.R., E.A., M.Z., R.A., H.G., P.N., N.C, M.Z., J.G.G., D.G.C., D. P., A.R., I.S.A., G.O., A.E.F. Writing-original draft: S.E., C.D., C.L., K.Z., R.Q.L, I.K., D.O., J.S., K.M., B.V., D.F.; Writing-review and editing: all authors.

