## Supplemental Information for "Biallelic pathogenic variants in TRMT1 disrupt tRNA modification and induce a syndromic neurodevelopmental disorder"

**Supplementary information**

**Efthymiou et al.**

Prenatal/Neonatal Course and Presenting Complaints (available upon request)

Supplemental clinical case reports (available upon request)

Supplementary Figure 1. Brain MRI plot

Supplementary Figure 2. Agarose gel of splicing analysis

Supplementary Figure 3. Primer extension analysis

Supplementary Figure 4. Zebrafish behavioral analyses

Supplementary Table 1. TRMT1 variants (available upon request)

Supplementary Table 2. Extensive clinical table of TRMT1 patients (available upon request)

Supplementary Table 3. Dysmorphology details (available upon request)

Supplementary Table 4. Splice predictions of *TRMT1* variants

Supplementary Table 5. Overview of TA cloning and fragment analysis

Supplementary Table 6. Fragment analysis

Supplementary Table 7. Minigene assay primers

Supplementary Table 8. Zebrafish primers

Supplementary Video 1. Patient 1 from family 1, available upon request

Supplementary Video 2. Patient 2 from family 1, available upon request

**Prenatal/Neonatal Course and Presenting Complaints**

Please contact corresponding authors for data.

**Supplemental clinical case reports**

Please contact corresponding authors for data.

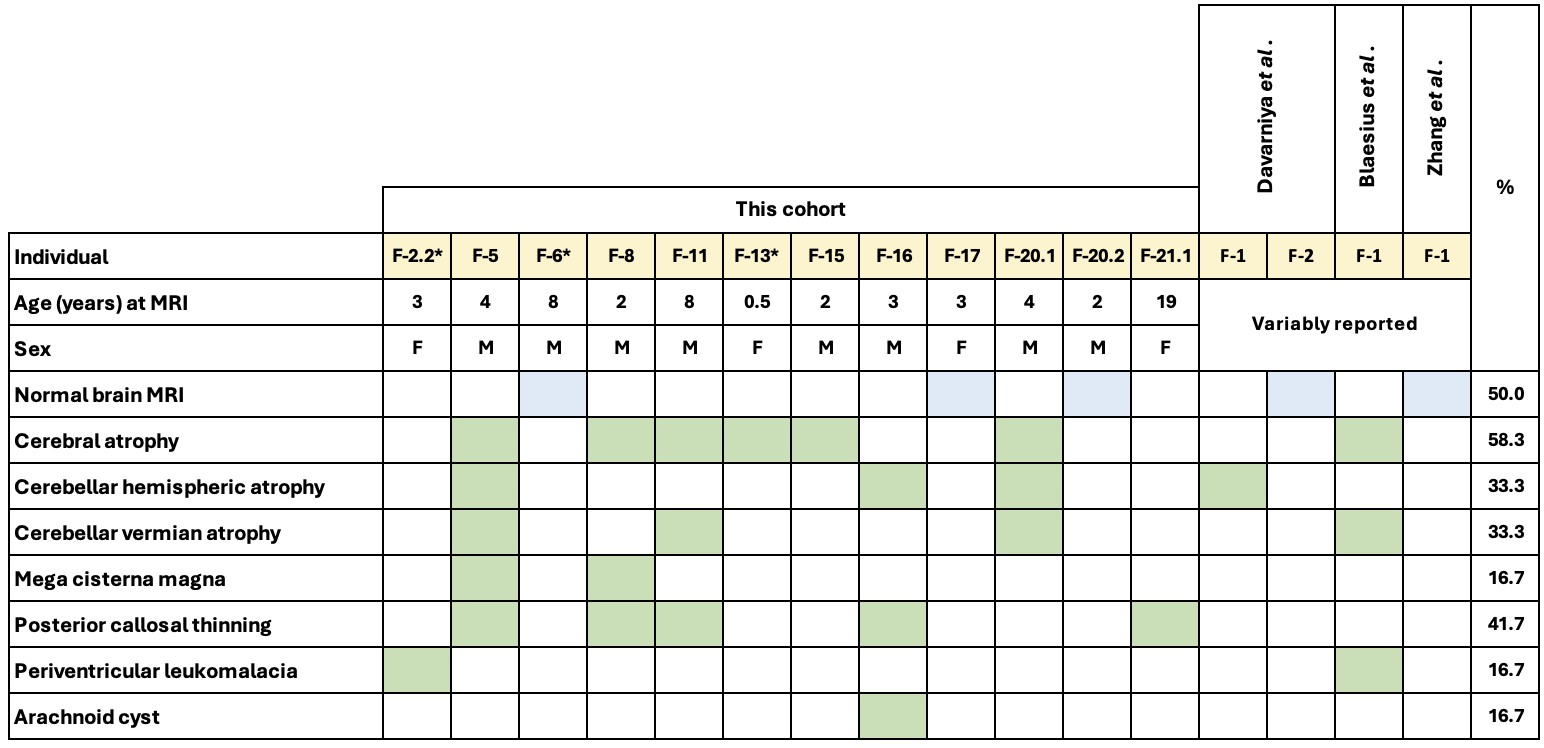

**Supplementary Figure 1. Neuroimaging features of *TRMT1*-ID in our cohort (*n*=12) and previously reported cases (*n*=4) with neuroimaging available for review.** Despite significant phenotypic heterogeneity, the most prevalent neuroimaging features were cerebral and cerebellar atrophy, the latter of which could be restricted to either the vermis or cerebellar hemispheres. Thinning of the corpus callosum was typically limited to the isthmus and splenium, with thinning of the callosal body present in a minority and uniform thinning present in only one individual.

100 bp ladder

c.310+5G>C

c.311-1G>A

c.454-1G>C

construct 1 WT

c.255-1G>T

construct 2 WT

c.1194G>A

construct 3 WT

Empty vector

c.1107-1G>A

Transfection negative

PCR negative

Construct 1 (ex3-5)

Construct 2

(ex9-10)

Construct 3

(ex11)

bp

100

200

300

500

400

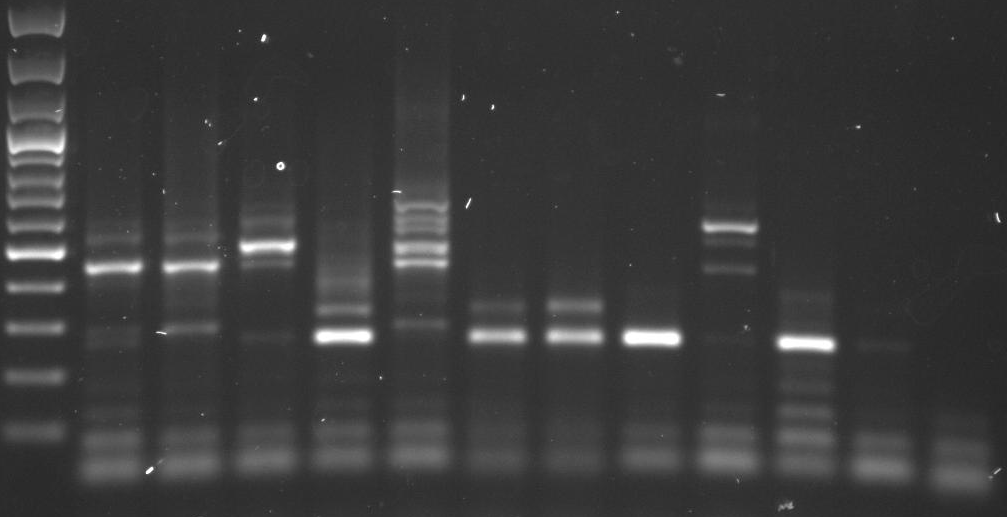

600

1

**Supplementary Figure 2. Agarose gel analysis of RT-PCR reactions from *TRMT1* minigene reporter constructs.** The indicated constructs were transfected into 293T human cells and RT-PCR performed on RNA. The reactions were loaded onto a 1% agarose gel and visualized by staining.

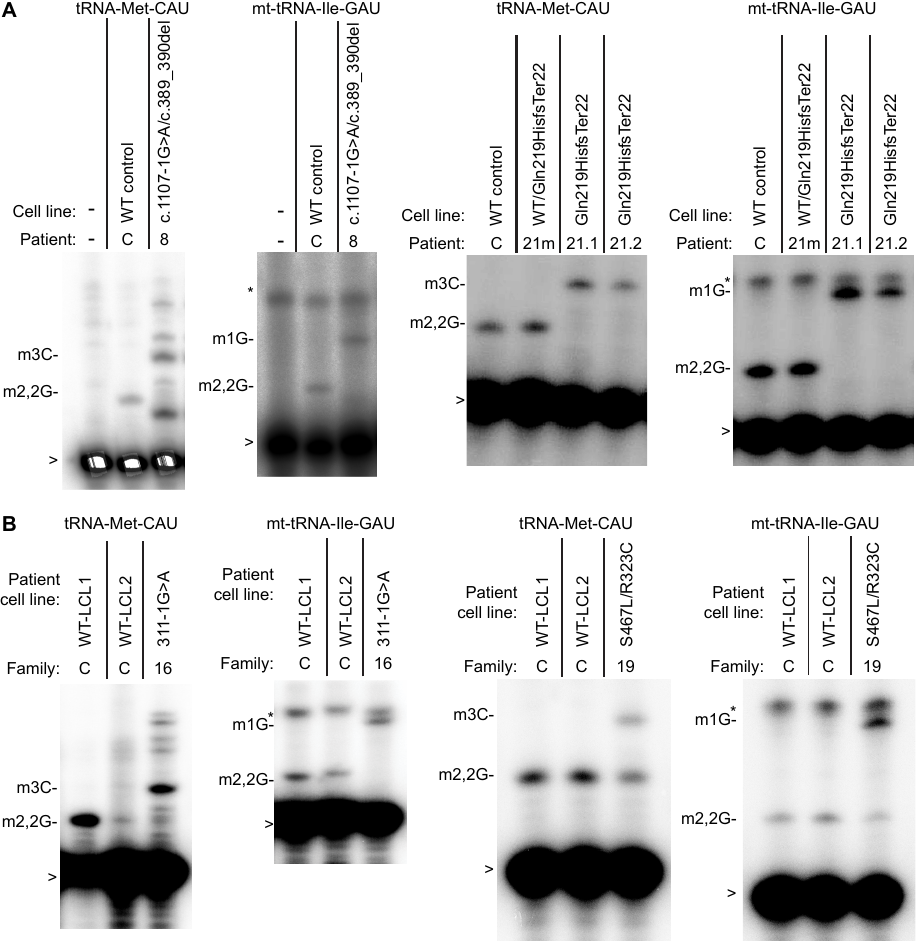

**Supplementary Figure 3. Patient cells with biallelic *TRMT1* variants exhibit a reduction in m2,2G modifications in tRNAs.** Representative gels of primer extension assays to monitor the presence of m2,2G in tRNA-Met-CAU and mt-tRNA-Ile-GAU from: (A) fibroblast cell lines or (B) lymphoblastoid cell lines. m3C_20_, 3‐methylcytosine; m2,2G_26_, dimethylguanosine; m1G_9_, 1‐methylguanosine; >, labeled oligonucleotide used for primer extension; *, background signal.

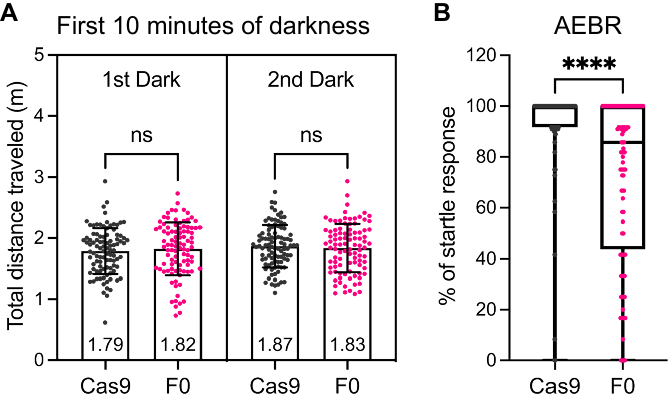

**Supplementary Figure 4. Behavioural analysis of control Cas9-injected and *trmt1* F0 knockout animals.** (**A**) Total distance traveled of each larva in the first 10 minutes of dark cycles. (**B**) A box and whisker plots showed *trmt1* F0 knockout larvae have less response to the sound stimuli. Statistical significances were calculated by unpaired *t* test with Welch’s correction: not significant (ns) *p* ≥ 0.05, ****p* < 0.001, and *****p* < 0.0001.
