## Supplementary Data for "Biallelic pathogenic variants in TRMT1 disrupt tRNA modification and induce a syndromic neurodevelopmental disorder"

### Slide 1
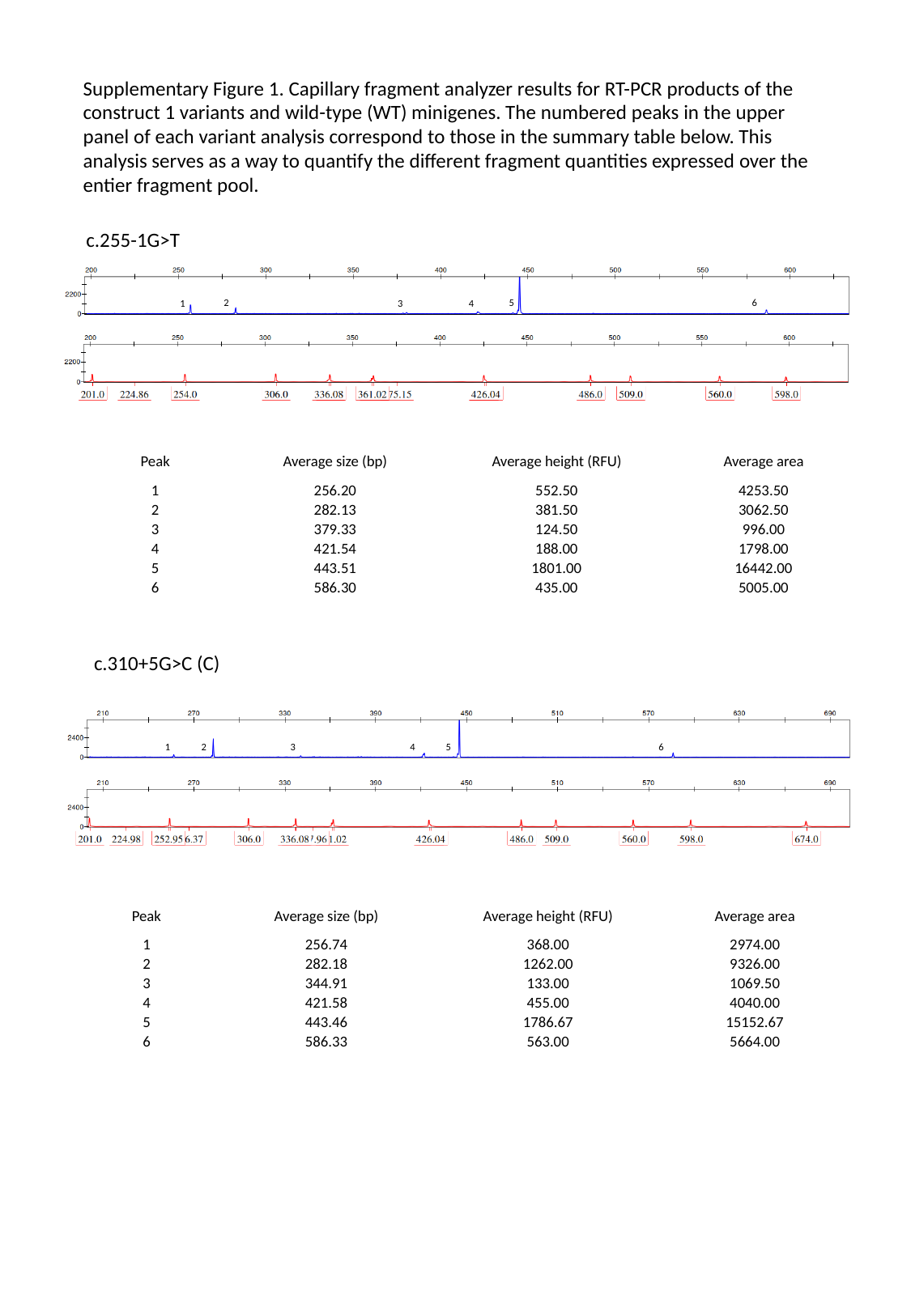

Supplementary Figure 1. Capillary fragment analyzer results for RT-PCR products of the construct 1 variants and wild-type (WT) minigenes. The numbered peaks in the upper panel of each variant analysis correspond to those in the summary table below. This analysis serves as a way to quantify the different fragment quantities expressed over the entier fragment pool.
c.255-1G>T
2
5
6
1
3
4
| Peak | Average size (bp) | Average height (RFU) | Average area |
| --- | --- | --- | --- |
| 1 | 256.20 | 552.50 | 4253.50 |
| 2 | 282.13 | 381.50 | 3062.50 |
| 3 | 379.33 | 124.50 | 996.00 |
| 4 | 421.54 | 188.00 | 1798.00 |
| 5 | 443.51 | 1801.00 | 16442.00 |
| 6 | 586.30 | 435.00 | 5005.00 |
c.310+5G>C (C)
3
1
2
4
5
6
| Peak | Average size (bp) | Average height (RFU) | Average area |
| --- | --- | --- | --- |
| 1 | 256.74 | 368.00 | 2974.00 |
| 2 | 282.18 | 1262.00 | 9326.00 |
| 3 | 344.91 | 133.00 | 1069.50 |
| 4 | 421.58 | 455.00 | 4040.00 |
| 5 | 443.46 | 1786.67 | 15152.67 |
| 6 | 586.33 | 563.00 | 5664.00 |

### Slide 2
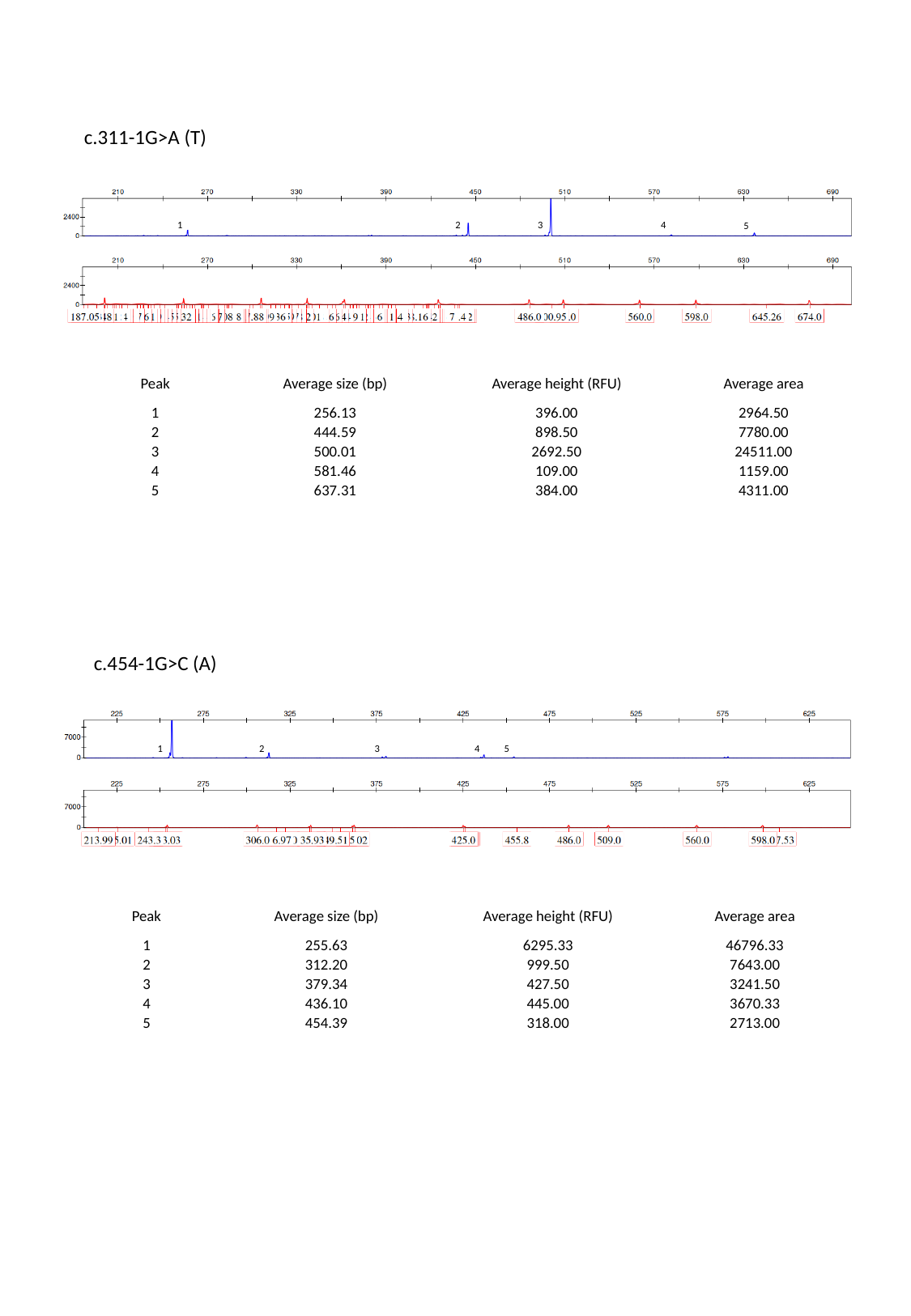

c.311-1G>A (T)
1
2
3
4
5
| Peak | Average size (bp) | Average height (RFU) | Average area |
| --- | --- | --- | --- |
| 1 | 256.13 | 396.00 | 2964.50 |
| 2 | 444.59 | 898.50 | 7780.00 |
| 3 | 500.01 | 2692.50 | 24511.00 |
| 4 | 581.46 | 109.00 | 1159.00 |
| 5 | 637.31 | 384.00 | 4311.00 |
c.454-1G>C (A)
1
2
3
4
5
| Peak | Average size (bp) | Average height (RFU) | Average area |
| --- | --- | --- | --- |
| 1 | 255.63 | 6295.33 | 46796.33 |
| 2 | 312.20 | 999.50 | 7643.00 |
| 3 | 379.34 | 427.50 | 3241.50 |
| 4 | 436.10 | 445.00 | 3670.33 |
| 5 | 454.39 | 318.00 | 2713.00 |

### Slide 3
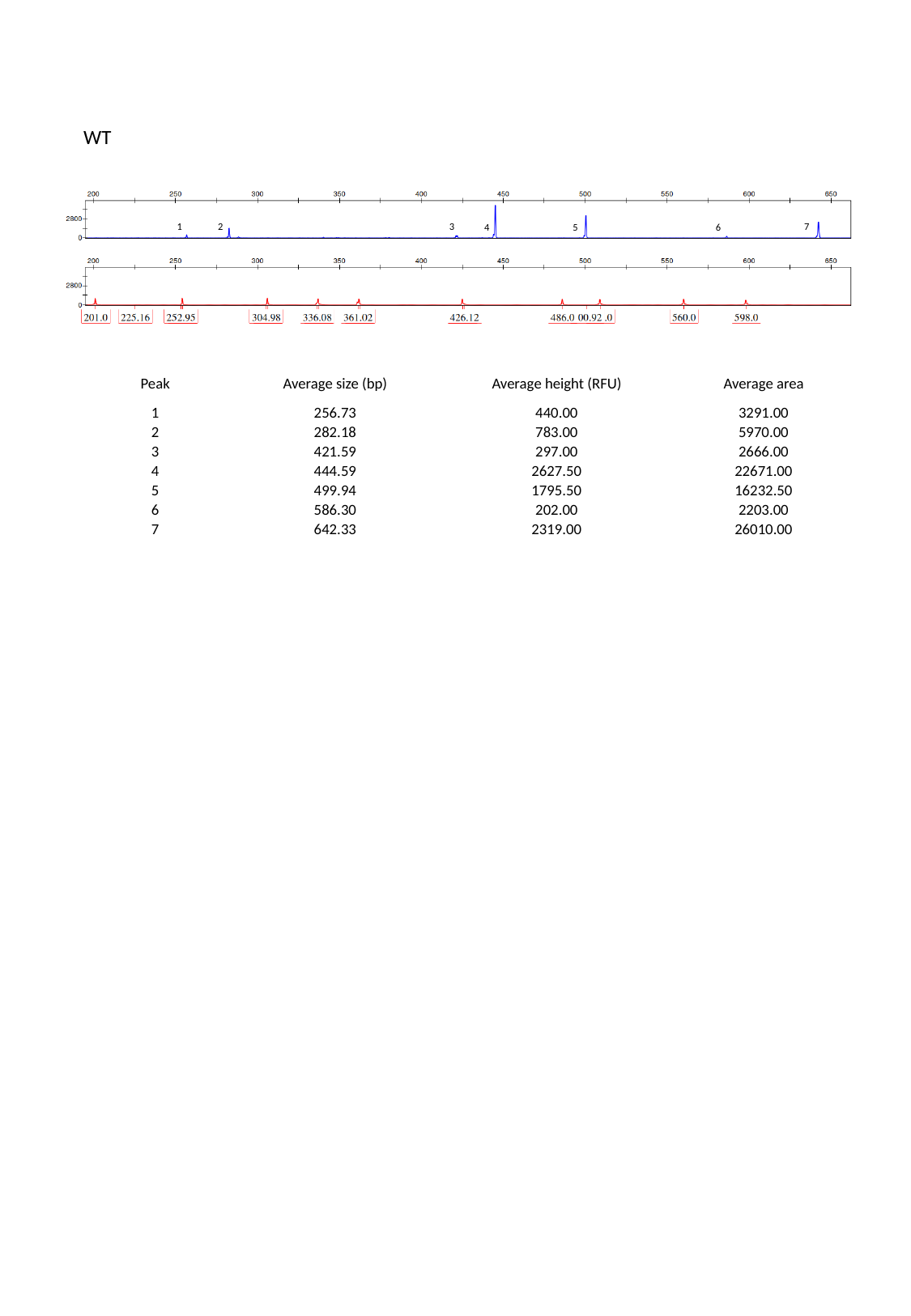

WT
7
1
2
3
4
5
6
| Peak | Average size (bp) | Average height (RFU) | Average area |
| --- | --- | --- | --- |
| 1 | 256.73 | 440.00 | 3291.00 |
| 2 | 282.18 | 783.00 | 5970.00 |
| 3 | 421.59 | 297.00 | 2666.00 |
| 4 | 444.59 | 2627.50 | 22671.00 |
| 5 | 499.94 | 1795.50 | 16232.50 |
| 6 | 586.30 | 202.00 | 2203.00 |
| 7 | 642.33 | 2319.00 | 26010.00 |
