## Supplementary Table 4 for "Biallelic pathogenic variants in TRMT1 disrupt tRNA modification and induce a syndromic neurodevelopmental disorder"

**Supplementary Table 4.** Splice Predictions of *TRMT1* variants

| Prediction Tool | c.255-1G>T | c.310+5G>C | c.311-1G>A | c.454-1G>C | c.1107-1G>A^1^ | c.1194G>A^2^ |
| --- | --- | --- | --- | --- | --- | --- |
| SpliceSiteFinder-like | -100% | -100% | -100% | -100% | -100% | +77.2% |
| MaxEntScan | -100% | -100% | -100% | -100% | -100% | +35.8% |
| NNSPLICE | -100% | -98.9% | No prediction | -100% | -100% | +90.0% |
| GeneSplicer | -100% | -100% | -100% | -100% | -100% | +9.2% |
| SpliceAI 10k  [≥0.2\|0.5\|0.8] | 0.79 AL (-1) | 0.92 DL (5) | 0.77 AG (-6)  0.90 AL (-1) | 0.50 AG (-5)  0.99 AL (-1) | 0.85 AG (-2)  0.94 AL (-1) | No change |
| AbSplice  [≥0.01\|0.05\|0.2] | 0.32 | 0.31 | 0.11 | 0.37 | 0.30 | No change |

Abbreviations: AG, acceptor gain; AL, acceptor loss; DL, donor loss

^1^Acceptor loss relative to native splice acceptor site

^2^Cryptic donor gain
