## Supplementary Table 5 for "Biallelic pathogenic variants in TRMT1 disrupt tRNA modification and induce a syndromic neurodevelopmental disorder"

**Supplementary Table 5.** Overview of percentage of TA cloning (TA) and Fragment analysis (FA) amplicons

| **Prediction Tool** | **c.255-1G>T** | | **c.310+5G>C** | | **c.311-1G>A** | | **c.454-1G>C** | | **WT** | |
| --- | --- | --- | --- | --- | --- | --- | --- | --- | --- | --- |
|  | **TA** | **FA** | **TA** | **FA** | **TA** | **FA** | **TA** | **FA** | **TA** | **FA** |
| **Exon 3 + Exon 4 + Exon 5 (644 bp)*** |  |  |  |  |  | 10.6 |  |  | 7 | 35.6 |
| **Exon 4 + Exon 5 (588 bp)** |  | 17.6 |  | 20.3 |  | 2.9 |  |  |  | 3.0 |
| **Exon 3 + Exon 4 + short Exon 5 (566 bp)** |  |  |  |  |  |  | 3.3 |  | 1.7 |  |
| **Exon 3 + Exon 5 (501 bp)** | 3 |  |  |  | 87.5 | 60.2 |  |  | 28 | 22.2 |
| **Exon 5 (445 bp)*** | 87.5 | 57.7 | 69 | 54.5 |  | 19.1 |  | 4.2 | 58 | 31.0 |
| **Exon 3 + short Exon 5 (423 bp)** |  | 6.3 |  | 14.5 |  |  | 3.3 | 5.7 | 1.7 | 3.7 |
| **Short Exon 5 (381 bp)** | 9 | 3.5 | 31 |  |  |  | 3.3 | 5.1 |  |  |
| **Exon 3 (313 bp)** |  |  |  |  |  |  | 23.3 | 12 |  |  |
| **All Exons skipped**  r.255_641del, p.Cys86Glnfs*24 |  | 14.9 |  | 10.7 | 12.5 | 7.2 | 66.6 | 73 | 3.5 | 4.5 |

*These Exons are present in protein coding transcripts.
