## Supplementary Table 6 for "Biallelic pathogenic variants in TRMT1 disrupt tRNA modification and induce a syndromic neurodevelopmental disorder"

**Supplementary Table 6.** Fragment analysis-based quantitative analysis of splice products from minigenes.

*TRMT1* c.255-1G>T

| **Average size (bp)** | **Actual size (bp)** | **Fragment (on gel)** | **Detected on gel** | **Average area** | **Percentage** |
| --- | --- | --- | --- | --- | --- |
| 586 | 588 | - | N | 5005.00 | 15.8 |
| 444 | 445 | 5 | Y | 16442.00 | 52.1 |
| 422 | 423 | - | N | 1798.00 | 5.7 |
| 379 | 381 | - | N | 996.00 | 3.2 |
| 282 | n.a. | - | N | 3062.50 | 9.7 |
| 256 | 257 | 9 | Y | 4253.50 | 13.5 |

| **Average size (bp)** | **Actual size (bp)** | **Fragment (on gel)** | **Detected on gel** | **Average area** | **Percentage** |
| --- | --- | --- | --- | --- | --- |
| 586 | 588 | - | N | 5664.00 | 14.8 |
| 443 | 445 | 5 | Y | 15152.67 | 39.6 |
| 422 | 423 | - | N | 4040.00 | 10.6 |
| 345 | n.a. | - | N | 1069.50 | 2.8 |
| 282 | n.a. | + | Y | 9326.00 | 24.4 |
| 257 | 257 | 9 | Y | 2974.00 | 7.8 |

*TRMT1* c.310+5G>C

A plus symbol denotes a band unable to be characterized from fragment analysis

*TRMT1* c.311-1G>A

| **Average size (bp)** | **Actual size (bp)** | **Fragment (on gel)** | **Detected on gel** | **Average area** | **Percentage** |
| --- | --- | --- | --- | --- | --- |
| 637 | 644bp | 1 | Y | 4311.00 | 10.6 |
| 581 | 588 | 2 | Y | 1159.00 | 2.9 |
| 500 | 501 | 4 | Y | 24511.00 | 60.2 |
| 445 | 445 | 5 | Y | 7780.00 | 19.1 |
| 256 | 257 | 9 | Y | 2964.50 | 7.2 |

| *TRMT1* c.454-1G>C |
| --- |

| **Average size (bp)** | **Actual size (bp)** | **Fragment (on gel)** | **Detected on gel** | **Average area** | **Percentage** |
| --- | --- | --- | --- | --- | --- |
| 454 | 445 | 5 | Y | 2713,00 | 4.2 |
| 436 | 437 | 6 | Y | 3670,33 | 5.7 |
| 379 | 381 | 7 | Y | 3241,50 | 5.1 |
| 312 | 313 | - | N | 7643,00 | 12 |
| 256 | 257 | 9 | Y | 46796,33 | 73 |

*TRMT1* WT

| **Average size (bp)** | **Actual size (bp)** | **Fragment (on gel)** | **Detected on gel** | **Average area** | **Percentage** |
| --- | --- | --- | --- | --- | --- |
| 642 | 644 | 1 | Y | 26010.00 | 32.9 |
| 586 | 588 | 2 | Y | 2203.00 | 2.8 |
| 500 | 501 | 4 | Y | 16232.50 | 20.5 |
| 445 | 445 | 5 | Y | 22671.00 | 28.7 |
| 422 | 423 | - | N | 2666.00 | 3.4 |
| 282 | n.a. | - | N | 5970.00 | 7.5 |
| 257 | 257 | 9 | Y | 3291.00 | 4.2 |
