## Supplementary Table 7 for "Biallelic pathogenic variants in TRMT1 disrupt tRNA modification and induce a syndromic neurodevelopmental disorder"

**Supplementary Table 7.** Minigene assay primers

| Region of Interest | Primer Name | Primer Sequence 5´- 3´ | Product Size |
| --- | --- | --- | --- |
| Construct 1  Exons 3-5 | TRMT1 Ex3-5 XhoI F | aattctcgagGAATTCAATCGGGACCTGAC | 1002 bp |
|  | TRMT1 Ex3-5 BamHI R | attggatccGGGCTCAAAGAGGCTAAGTC |  |
| Construct 2  Exons 9-10 | TRMT1 Ex9-10 XhoI F | aattctcgagCCAGTCTAAGGGAGGAGTTGG | 416 bp |
|  | TRMT1 Ex9-10 BamHI R | attggatccGATGGGCTCTGCCCACAT |  |
| Construct 3  Exons 11-12 | TRMT1 Ex11-12 EcoRI F | aattGAATTCTTAGGGCCAAGTTCTCTGCA | 446 bp |
|  | TRMT1 Ex11-12 NotI R | attGCGGCCGCTGGTGTGTTGCAGTGGATG |  |
| pSPL3 Exons A and B | SD6 F | TCTGAGTCACCTGGACAACC | -- |
|  | SA2 R | ATCTCAGTGGTATTTGTGAGC |  |
| pSPL3 Exons A and B | SD6 F-FAM | FAM-TCTGAGTCACCTGGACAACC | -- |
|  | SA2 R-FAM | FAM-ATCTCAGTGGTATTTGTGAGC |  |
| Vector pCR2.1 | M13 F | GTAAAACGACGGCCAG | -- |
|  | M13 R | CAGGAAACAGCTATGACC |  |
