## Supplementary Table 8 for "Biallelic pathogenic variants in TRMT1 disrupt tRNA modification and induce a syndromic neurodevelopmental disorder"

**Supplementary Table 8.** Sequences of sgRNAs and RT-qPCR primers for zebrafish experiments.

| All sequences are given in 5' to 3' direction | |
| --- | --- |
| **sgRNA target sequence for generating F_0_ knockout** | |
| trmt1_gRNA_1 | GTGTAGGATCTTCCAGTGTG |
| trmt1_gRNA_2 | GGCATGATCTTCAGTATGTG |
| trmt1_gRNA_3 | GTGTAGGATCTTCCAGTGTG |
| **Primers used for RT-qPCR** | |
| trmt1_set-1_Forward | TGCTCTGGAAGTTCCTGGCC |
| trmt1_set-1_Reverse | GATGACATCATAGCGCTCCTTCC |
| trmt1_set-2_Forward | GGGTGTTATGGGATATCATGCGC |
| trmt1_set-2_Reverse | CTCTTGTCCTCGAGCTCTGAGG |
| 18S_Forward | TCGCTAGTTGGCATCGTTTATG |
| 18S_Reverse | CGGAGGTTCGAAGACGATCA |
